# Co-existence of negative and positive associations between cognition and intergenerational psychiatric symptoms reveal necessity of socioeconomic and clinical enrichment

**DOI:** 10.1101/2023.08.28.23294743

**Authors:** Adam Pines, Leonardo Tozzi, Claire Bertrand, Arielle S. Keller, Xue Zhang, Susan Whitfield-Gabrieli, Trevor Hastie, Bart Larsen, John Leikauf, Leanne M. Williams

## Abstract

**Background:** Mental illnesses are a leading cause of disability globally. Across 17 psychiatric disorders, functional disability is often in part caused by cognitive impairments. However, cognitive heterogeneity in mental health is poorly understood, particularly in children.

**Methods:** We used generalized additive models (GAMs) to reconcile discrepant reports of cognitive impairment across classes of psychiatric symptoms in 4,782 children and their parents. Specifically, we derive relationships between cognition and psychopathology across different ranges and classes of symptom burdens. We additionally evaluate generalizability across sex-assigned-at-birth (SAAB) and federal poverty status. Finally, we incorporate a measure of scholastic performance as a real-world measure of functional ability. Associations were tested at the 99% confidence level.

**Results:** We demonstrate that the previously-reported, weak, negative, and linear relationship between general cognition and general psychopathology consists of several stronger but opposed relationships. Externalizing symptoms are negatively associated with cognition, but internalizing symptoms are positively associated with cognition at low symptom burdens. This phenomenon holds across parental and child symptoms. Finally, we provide evidence that, compared to laboratory measures of cognition, school grades are more accurate and generalizable indicators of psychopathological burden in children.

**Discussion:** The most common approach to quantifying the relationship between cognition and psychopathology systematically underestimates the strength and complexity of this relationship. Grades may represent a more accurate and generalizable marker of mental illness. Developmental studies incorporating clinical enrichment, parental mental health, and socioeconomically diverse samples may provide deeper and more generalizable insight into neurocognitive impairment and psychopathology.

## Introduction

Depression, anxiety, and behavioral disorders are among the leading causes of illness and disability in adults^1^ and adolescents^2^. Study of cognitive impairment is paramount to understanding these disabilities: cognitive impairments are near-ubiquitous across psychiatric disorders, constituting diagnostic criteria for 17 different conditions^3^. Further, in adults, cognitive impairments are often responsible for impaired daily functions in psychiatric conditions, which are in turn directly associated with decreased quality of life^4,5^. Because of the ubiquity and malignancy of cognitive impairments across psychiatric disorders, cognitive impairment may constitute a critical transdiagnostic phenotype in its own right^6–8^.

Convergent psychiatric, neuroimaging, and animal studies have implicated cognitive impairment in mental illness^9^, but cognitive deficits and the daily functions they impair manifest heterogeneously across individuals^10,11^. Recent evidence suggests that neurocognitive variability might be amplified in children^12–14^ and linked to latent psychiatric risk^15,16^. This inter-child heterogeneity may arise from heightened environmental and familial susceptibility^17–19^, which affords sociobiological factors greater influence to directly give rise to psychiatric symptoms in children (such as the financial means^20,21^ and mental health of ones’ family^22,23^). These sociobiological factors are often intergenerational^24,25^ and sex-dependent^26,27^, further complicating inter-child heterogeneity in cognitive functions^18,28–30^. This heterogeneity can frustrate efforts to derive generalizable and actionable patterns of psychiatric impairment in children^31,32^. Preliminary evidence in German^5^ and Chilean^33^ schoolchildren does suggest that scholastic grades may constitute a generalizable link between cognitive impairment and mental health. Due to elevated vulnerability and variability in children, delineating the generalizability of scholastic-psychiatric associations across sociodemographics is imperative to understanding how cognition is linked to mental health in youth.

However, a critical obstacle in understanding cognitive impairment in psychopathology is that the link between cognition and psychopathology is itself contentious. Recent studies, often in large samples, have reported both negative^34–38^ and positive^39–43^ associations between cognition and symptom burdens. Although these studies have evaluated varying levels of symptoms (from healthy participants to extremely unwell), the use of linear modeling has precluded such studies from determining whether the association between cognition and psychopathology varies across different symptom burdens. Linear approaches implicitly assume a one-size-fits-all, fixed relationship (slope) between cognition and psychopathology across all ranges of symptom burdens. Although linear approaches are intuitive, robust, and often better-suited for small sample sizes, they cannot capture instances where both excessively high or low measurements of a clinical variable are indicative of clinical risk, such as weight, blood pressure, or time spent sleeping. Because prior studies have investigated cognition and psychopathology across heterogeneous ranges of clinical symptoms, and because linear approaches implicitly assume the same fixed slope irrespective of these ranges, it is possible that seemingly conflicting reports may arise purely from differences in the ranges of symptoms studied. In other words, reports of both positive and negative associations between symptoms and cognition can simultaneously be true, if the association depends on the level of symptom burden.

Recently, generalized additive models (GAMs) have been successfully leveraged to accurately delineate complex, nonlinear relationships between cognitive, developmental, and physiological variables of interest^44–48^. Through explicit mitigation of overfitting, these models afford a rigorous approach to decomposing nonlinear relationships between complex variables of interest^49,50^. Critically, GAMs allow assessment of continuous relationships between risk factors and health outcomes when both high and low measurements indicate clinical risk. Furthermore, such models can account for continuous relationships between clinical variables and thus may more accurately link naturalistic variation in cognition to individual differences in mental illness symptoms than linear or categorical models of cognitive dysfunction.

Here, we use a large, diverse, and multi-generational sample (Adolescent Brain Cognitive Development Study®, ABCD^51,52^) to demonstrate that relationships between cognition and symptom burden are both negative and positive, depending on several key factors. We provide evidence that the commonly-reported weak linear relationship between symptom burden and cognition consists of several stronger-but-conflicting relationships for different symptoms in children. By characterizing inter-individual heterogeneity across multiple sociobiological factors, we find that the strength and direction of the association between cognition and symptom burden depend on the class of symptoms, the range and magnitude of symptom burden, parental mental health, sex-assigned-at-birth (SAAB), and whether or not children live in poverty. Finally, in light of this complexity, we demonstrate that scholastic grades may be a more accurate and generalizable indicator of psychopathological burden in children.

## Methods

### Study design and participants

Data were drawn from the Adolescent Brain Cognitive Development study (ABCD®), collected across 21 sites in the United States. Measurements were conducted at two timepoints, with the first occurring when the child is 9 to 11 years of age (mean age = 9.9 years old, *S.D.* = 0.6 years) and the second occurring two years later (mean age = 11.9, *S.D.* = 0.6 years). Exclusion criteria for participation in the ABCD study included lack of child and parental fluency in English, major medical conditions, past traumatic brain injuries or other contraindications to MRI scanning, and pre-term birth (gestational age <28 weeks).

As we aggregated our measurements of interest, additional exclusions were incurred when participant-level data was missing for variables of interest (**Figure S1**). However, comparison of initial and final sample suggested that missing data did not systematically exclude any race or ethnicity of children. Our final sample was comprised of 42.5% ethnic minority groups, which compares favorably with the composition of the United states (41.1%^53^) as well as the United Nations’ estimate of 10-20% of individuals belonging to ethnic minority groups globally^54^. However, Asian Americans were disproportionately underrepresented in our initial and final sample (2% of sample prior to and after excluding missing data). Our aforementioned measures of interest are detailed below.

### Cognitive measures

Cognitive measures were drawn from the National Institutes of Health (NIH) Toolbox and several complementary cognitive tasks (Rey Auditory Verbal Learning and Delayed Recall tests, Matrix Reasoning, and the Little Man task)^55^. To maximize the reliability of our findings, we chose to measure cognitive performance across tasks with a single general cognition (*g*) factor. The *g* factor exhibits strong generalizability across diverse populations^56^, and is reliably delineated across independent cognitive batteries^57^. We modeled our approach after Thompson et al., 2019^19^. We used principal component analysis (PCA) to extract a single *g* factor as the component explaining the most variance across cognitive tasks that were available at both timepoints. We demonstrate that this process yields an equivalent factorization to that derived from Bayesian PCA in Thompson et al. (*r =* 0.98, *p* < 0.0001, **Figure S2**). The measures included in Thompson et al. but not available at timepoint two were the list sorting task (72 total values included in timepoint two data frame) and the card sorting task (65 total values included in timepoint two data frame). PCA was conducted after retaining a single child from each family, so that every family was equally represented in the delineation of *g* and subsequent results.

### Symptom measures of psychopathological burden: children

Child psychopathology was evaluated from the Child Behavioral Checklist (CBCL)^58^. The CBCL assesses a wide range of symptoms and gauges the degree to which any given symptom is experienced. For each symptom, parents/guardians can report either 0 for “Not True”, 1 for “Somewhat or sometimes true” or 2 for “Very true or often true” with respect to their child. Parent-reported child symptoms were utilized due to evidence of a stronger correspondence between clinician evaluation and parental report than clinician evaluation and child self-report^59^. Similarly to general cognition (*g*), we utilize a general psychopathology factor (*p)* score. Specifically, we use the raw sum of endorsed symptoms, with a maximal value of 2 (“Very true or often true”) for each value. Raw sum was chosen over alternative derivations of *p* due to improved model fits and interpretive value, as detailed below.

First, to ensure the raw sum of endorsed symptoms produced a comparable measurement to formal *p* factor derivations, we replicated a PCA-derived *p* factor score in accordance with Michelini et al., 2019^60^. Specifically, we inverted values on reverse-scored items and removed collinear items (polychoric r > 0.75) prior to PCA, from which the first component was extracted as a general *p* factor. Because these formal *p* factor values were derived from principal component analysis, this approach transformed counts of endorsed symptoms into a continuum of negative-to-positive loadings. Negative values, intended to represent the degree to which symptoms are experienced, presented interpretative difficulties. Further, in contrast to cognitive scores, symptom endorsements were heavily skewed (Lilliefors test for normality in baseline, 2-year data: D = 0.15, 0.16, *ps ≈ 0*). Although link functions exist for regressions when conditional response variables are heavily skewed, many of these link functions are not compatible with negative data (i.e., negative binomial). Consequently, in instances where we modeled psychopathology as a response variable, this transformation of PCA prevented us from optimally modeling psychopathology outcomes. Specifically, fitting PCA-derived *p* as a response variable with *g* as a predictor yielded systematic overprediction of *p* in children with low *g* values and systematic underprediction of *p* in children with high *g* values (**Figure S3**, left). In contrast to the standard gaussian link function, a negative binomial link function, explicitly intended for positive and non-normally distributed count data (rather than mixed positive and negative values), fit this relationship substantially better (**Figure S3**, right). Further, raw count of endorsed symptoms was highly correlated (*r* = 0.99, *p* < 0.001) with formal PCA-derived *p* factors, suggesting that the raw symptom count is functionally equivalent to PCA-derived *p*-factor values (**Figure S4**). Because raw symptom counts resulted in models that were less systematically biased against children with low and high symptom scores (**Figure S3**) and because formal *p*-factor scores appeared to be functionally equivalent to raw symptom counts (**Figure S4**), we proceeded with raw symptom count (total CBCL score) as our *p* factor for children.

To capture dimensional variation across classes of psychiatric symptoms, we also extracted raw symptom counts across CBCL subscales. CBCL subscales include aggression, anxious depression, attention, rule breaking, social, somatic, thought, and withdrawn depression. Subscales also include externalizing symptoms, which is the sum of both aggressive and rule breaking behaviors, as well as internalizing symptoms, which is a sum of anxious depression, somatic, and withdrawn depression subscales.

### Clinical thresholds

To contextualize different magnitudes of reported symptoms, we included vertical dashed lines at borderline-clinical and clinical thresholds in plots depicting CBCL scores. Borderline-clinical thresholds and clinical thresholds were calculated from *t-* scores and translated to raw scores. Specifically, we used the established *t-*score for borderline clinical thresholds and clinical thresholds (*t* = 65-69 used for borderline-clinical range, *t* ≥ 70 for clinical range), and extracted the raw score matched to these *t*-scores (average raw score at *t*=65 used as borderline-clinical threshold, average raw score at *t*=70 used as clinical threshold). The symptom ranges under borderline-clinical thresholds are referred to as the subclinical range, and symptom ranges extending from clinical thresholds to the maximum symptom score sampled is referred to as the clinical range.

### Symptom measures of psychopathological burden: adults

Adult psychopathology was evaluated from the Adult Self Report (ASR) scale, which provides adult mental health measurements roughly equivalent to those derived from the CBCL. These items are self-reported by the parent/guardian, and also range from 0 (Not true) to 2 (Very true or often true). An equivalent general parental psychopathology score (parental *p*) was also derived as the sum of all items. Finally, we also included CBCL-equivalent ASR subscales for each class of symptoms, including aggression, anxious depression, attention, rule breaking, somatic, thought, and withdrawn depression, externalizing, and internalizing subscales. An equivalent social subscale was not measured in adults. To evaluate specific symptom ranges similarly to subclinical and clinical ranges for children, adult symptom ranges were also segmented into thirds based on the total range of symptoms sampled.

### Demographic and scholastic measures

Sex-assigned-at-birth (SAAB) was available for all participants, as reported by a caretaker. Child-identified gender was not available in the current data release (abcd_yksad01). Poverty status was defined as household income below 25,000 dollars (*n* = 617 children), given the federal poverty guidelines for a family of four from the time this data was acquired (< $25,100 household income per year^23^). This threshold aligned with the fourth income level queried in this study (<5k income, 5-12k, 12-16k, 16-25k, 25-35k, 35-50k, 50-75k, 75-100k, 100-200k, > 200k), which we derived from the ABCD-BIDS Community Collection (ABCC^61^).

Which children belong to which family was evaluated with the American Community Survey (ACS). This step was used to remove one sibling from families with multiple siblings enrolled, at random, from our dataset (**Figure S1**) to ensure that no family was over-represented in our factor extraction or results relative to other families.

Scholastic measurements at timepoint one were available through the Parent Diagnostic Interview for DSM-5 Background Items. These items provided a parent-reported measurement of grades considered average for the queried parent’s child (A’s, B’s, C’s, D’s, F’s, ungraded, or not applicable). At subsequent timepoints, grades were queried at finer-grained resolution through the ABCD Parent School Attendance and Grades questionnaire, which allows for specifying grade ranges within letter-grades (i.e., 80-82, 83-86, and 87-89 instead of a unitary “B” grade). To allow for equivalent analyses across timepoints, timepoint two grade measures were collapsed into broader letter-grade they fell into. Letter-grades were used for all scholastic analyses. In addition to children with missing scholastic data, children with ungraded or not applicable scholastic performance were not considered in our analyses.

### Psychopathological burden as a primary predictor

Throughout, we use the term ‘psychopathology’ to refer to CBCL subscales (or general psychopathology) from which psychopathological burden can be assessed continuously, from relative absence to clinically severe. Notably, ABCD included a wide range of healthy, subclinical, and clinical participants. To facilitate generalizability of our results across this entire range, we only present cognitive-symptom associations where we obtained at 99% or greater certainty of a positive or negative association (positive or negative slope in > 9,900 iterations of 10,000 bootstrap resamples).

### Cognition as a primary outcome

We have predominantly operationalized *g* as our dependent variable across timepoints in our analyses. The motivation for modeling *g* as an outcome is two-fold. First, as described, evidence suggests that impaired cognition may constitute a critical transdiagnostic clinical phenotype in its own right^6–8^, rather than being subsumed by the diagnosis of any particular patient. Second, although more timepoints may reveal bidirectional temporal precedence^62^, extant evidence and our own analyses (below) suggest that temporal precedence in the relationship between cognition and psychopathology is slightly more consistent with psychopathology preceding cognitive impairment than cognitive impairments preceding the development of symptoms in this age range^25,26^.

### Temporal precedence of predictor and outcome variables

In order to most ensure our choice of psychopathology as a predictor variable and cognition as an outcome variable was cohesive, we sought to replicate prior findings on temporal precedence between *g* and *p* in the ABCD study^63^. To do so, we deployed cross-lagged panel analysis with *lavaan* in R. This approach leverages longitudinal data to query if variables at one timepoint (here, *g* and *p* at timepoint 1) predict each other at another timepoint, above and beyond the starting values for those variables^64^. Autoregressive effects for *g*, *p*, Internalizing, and Externalizing symptoms were modeled in both timepoints, and cross-lagged effects were fit for *g* onto psychopathology variables and psychopathology variables onto *g*. We replicated extant evidence: from the two timepoints available, temporal precedence is generally more consistent with *p* predicting future *g* than *g* predicting future *p* (**Table S1**). Although psychopathology and cognition are likely both causal of each other^21,62,63^, our results suggest that timepoint one psychopathology variable explains more variance in timepoint two cognition variables than vice-versa in the current study.

### Statistical analyses

We used generalized additive models (GAMs) with penalized splines (denoted by s(x)) to simultaneously capture linear and non-linear relationships between cognition and psychiatric symptoms. Specifically, as prior, we fit generalized additive models with four knots per spline^47,48,65^. Because of the known positive association between age and cognitive scores in children, we also covaried for age as a spline in all analyses such that *g* ∼ s(age) + s(symptoms), where symptoms was the composite score (*p-*factor) or subscale scores. We implemented these models with the *bam* function *mgcv* package in R (version 4.1). To avoid overfitting, nonlinearity was penalized with restricted maximum likelihood^50^. Spline slopes were calculated by computing the derivative of the fitted spline functions.

To quantify significant positive and negative slopes across different levels of symptom burdens, both timepoints were merged prior to conducting 10,000 bootstrap resamples of our final participant sample. Subsequently, we calculated instances where levels of symptom burdens were consistently identified as either having a positive or negative slope in > 99% (> 9,900 iterations) of bootstrap resamples. Because we modeled data over two timepoints, bootstrap resamples were explicitly conducted to remove or retain entire *participants* from each resample rather than *observations*. In other words, if a participant was excluded from one bootstrap resample, data from neither of their timepoints was included. This procedure allowed us to account for potential inflation or underestimation of effects attributable to included participants. To avoid over-extrapolating our analyses, we limited our approach to symptom ranges represented in each bootstrap. Because conventional measurements of effect size are not coherent with generalized additive models (i.e., beta-weights reflect a fixed slope), we instead used deviance explained to capture effect sizes of comparable variables. Akaike information criterion (AIC) and deviance explained were also utilized to compare model fits.

To quantify generalizability, we constructed equivalent models within two sets of groups assembled on the basis of known sources of psychiatric heterogeneity: sex-assigned-at-birth (SAAB) and familial poverty. The association between general cognition and psychopathological burden was estimated across 10,000 bootstrap resamples across these groupings. Generalizability was operationalized as manifestation of the same direction of effect (positive vs. negative) over the same range of the independent (predictor) variable. Because there were fewer children within the familial poverty grouping (<$25,000 combined annual income, *n* = 617 vs. *n* = 4165), we conducted an additional analysis to ensure differential sample sizes were not underlying generalizability differences. Specifically, we created a size-matched non-poverty group in each bootstrap iteration, where the group size was constrained to be the same number of children meeting poverty criteria selected in any given bootstrap iteration.

### Openly-available analysis code

To facilitate transparency and reproducibility^66^, all utilized analysis code is available in the public domain at https://github.com/WilliamsPanLab/gp. Additionally, we have curated a detailed plain-language walk-through of all scripts utilized for analyses throughout this manuscript.

## Results

### The relationship between general cognition and psychopathology depends on class of symptoms and level of symptom burden

We first sought to re-establish the commonly-reported negative linear relationship between general cognition (*g*) and general psychopathology (*p*) in our current sample. In accordance with prior literature^35,67^, *g* and *p* were weakly negatively correlated across the full sample (*r* = −0.05, *p* < 0.001, *r*^2^= 0.003). However, comparison of *g*∼*p* with linear and spline fits for *p* suggested that the association between *g* and *p* is better-explained by a nonlinear relationship (**Table S2**). Generalized additive models revealed that the weak, aggregate relationship between *g* and *p* consisted of several stronger, but often opposed constituent relationships. First, the association between *g* and *p* varied by symptom severity: the negative association between *g* and *p* primarily manifested when symptom burden was reported at clinically-significant thresholds rather than evenly across the entire range of reported symptom (**Figure 1A**). Second, when *p* was decomposed into internalizing and externalizing symptoms, we found that the association between *g* and *p* varied by symptom class: we observed a negative association between *g* and externalizing symptoms, but negative *and* positive associations between *g* and internalizing symptoms across clinical and sub-clinical symptom burdens, respectively (**Figure 1B**).

**Figure 1:**
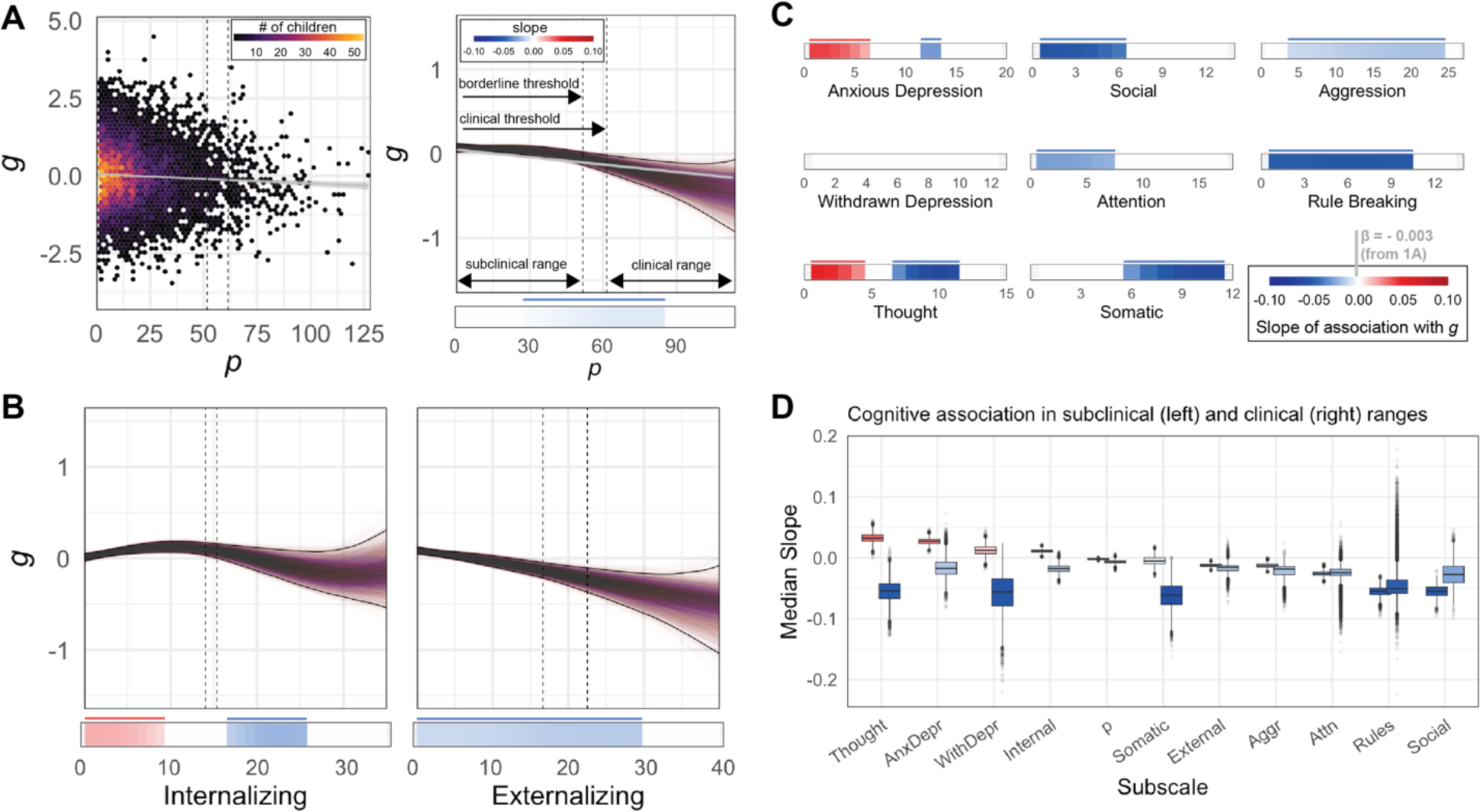
The relationship between general cognition and psychopathology consists of opposed, constituent relationships. **A)** Left: Consistent with prior literature, a linear model reveals that general cognition (*g*) is weakly, negatively associated with general psychopathology (*p*; dashed vertical lines = borderline and full clinical risk thresholds; *r* = - 0.05, *p* < 0.001, *r*^2^= −0.003). Right: Penalized splines in 10,000 bootstrap resamples reveal a non-linear relationship between *g* and *p*, such that *g* is more negatively associated with *p* at higher symptom burdens. Red and blue segments within basal horizontal bars depict where negative or positive slope was reliably detected across at least 99% of all bootstrap iterations. Specifically, each colored segment depicts where the 99% confidence intervals span exclusively negative (blue) or positive (red) first derivatives. **B)** Decomposition of general psychopathology into internalizing and externalizing symptoms reveals opposed, constituent cognitive associations. Internalizing symptoms are positively associated with cognition at low symptom burdens and negatively associated at clinical symptom burdens; externalizing symptoms only exhibit negative associations. **C)** Further decomposition of psychopathology into constituent subscales reveals stronger and often opposed relationships across different levels of symptom burden. Vertical gray bar reflects estimated linear slope (*r*^2^ = −0.003), from left panel in **1A**, is overlaid onto color bar for comparison to GAM-estimated slopes. **D)** All paired boxplots represent the median slope across bootstrap iterations for the subclinical (left) and clinical (right) range of symptoms for each CBCL scale (subclinical range vs. clinical range). In the subclinical range of symptoms, internalizing symptoms are positively associated with cognition. However, in the clinical range, symptoms are consistently negatively associated with cognitive symptoms. These findings suggest both the strength and complexity of the relationship between cognition and psychopathology is underestimated by aggregate linear models in children.

Further decomposition of endorsed symptoms into the other 8 subscales of the CBCL further revealed opposed constituent relationships underlying the association between *g* and *p*, such that more granular subscales of psychopathology were generally more strongly associated with *g* than coarser scales, often with different slopes at different levels of symptom severity (**Figure 1C**, full subscale plots in **Figure S5**). Comparison models suggested that nonlinear relationships between *g* and symptoms were more common for internalizing rather than externalizing subscales (**Table S2**). Full reversals, instances where symptoms were both negative and positively associated with *g* depending on symptom severities, were present for internalizing, anxious depression, and thought subscales (**Figure 1D**). These findings suggest concurrent positive and negative associations between cognition and psychopathology exist, and that the relationship between cognition and psychopathology depends on the class and severity of psychopathology.

Notably, variability in the correspondence between *g* and symptoms at different severities is incompatible with linear models assuming a fixed slope. As low levels of symptoms are not necessarily associated with concomitant cognitive impairments, this suggests that linear analyses assuming a one-size-fits-all slope will artifactually weaken relationships derived from a broad range of symptom severities. As proof of concept, we demonstrate that compared to the full sample, the effect size of the correlation between cognitive scores and symptoms is 8 times larger within the clinical range of the sample (*r*/*r*^2^ = −0.052/0.003 vs. *r*/*r*^2^ = −0.149/0.022), and 44 times larger than an effect size derived purely within the subclinical range of the sample (*r*/*r*^2^ = −0.023/0.0005 vs. *r*/*r*^2^ = - 0.149/0.022, **Figure S6**). These results reveal a clear dependency of linear, fixed-slope estimates between *g* and *p* on the range of psychopathology being assayed, such that correlational studies without substantial clinical enrichment are likely to considerably underestimate the negative relationship between *g* and clinical symptom burdens.

### Parental mental health is a dimension of inter-child variability in cognition

Cognition exhibited complex, multidirectional relationships with symptom burdens across domains of mental health. Importantly, mental health^68^ and cognitive abilities^69^ have strong intergenerational links^24,25^, and child development is strongly influenced by caregiver behaviors^18,24,70^. Consequently, understanding the link between parental mental health and child cognition is arguably as important as understanding the link between mental health and cognition within a child. To evaluate intergenerational contributors to general cognition in children across symptom classes and burdens in parental mental health, we evaluated parental mental health with the adult-adapted version of the CBCL (Adults Self-Report, ASR).

Unsurprisingly, we found a strong relationship between parental mental health and parent-reported child mental health (*r* = 0.57, *p* < 0.001, **Figure 2A**). As seen for internalizing symptoms for children, low-but-present levels of general psychopathology in parents were associated with higher child cognition, such that children with parents reporting just a few symptoms had higher cognitive scores than parents reporting minimal symptoms (**Figure 2A**). As for children, adult internalizing symptoms exhibited a non-linear association with child cognition (**Table S3**), such that the greatest negative associations with cognition were seen at higher symptom burdens. Contrastingly, adult externalizing symptoms were negatively associated with cognition across nearly the entire range of symptoms assayed (**Figure 2B**). The relationship between parental symptoms and child cognition also varied by domain of psychopathology: more granular parent symptom subscales revealed differential relations with child cognition (**Figure 2C, Figure S7**). Consistent with child symptoms (but inconsistent with assuming a fixed slope), the strongest negative association with cognition was observed at higher magnitudes of reported symptoms; rule-breaking behavior was the only exception to this pattern (**Figure 2C**). In addition to general psychopathology, parental thought disorders exhibited a clear reversal in the association with child cognition at different symptom burdens. Specifically, parental *p* and thought symptoms exhibited a positive association with *g* at low levels of symptoms, but a negative association at high symptom levels (**Figure 2D**).

**Figure 2:**
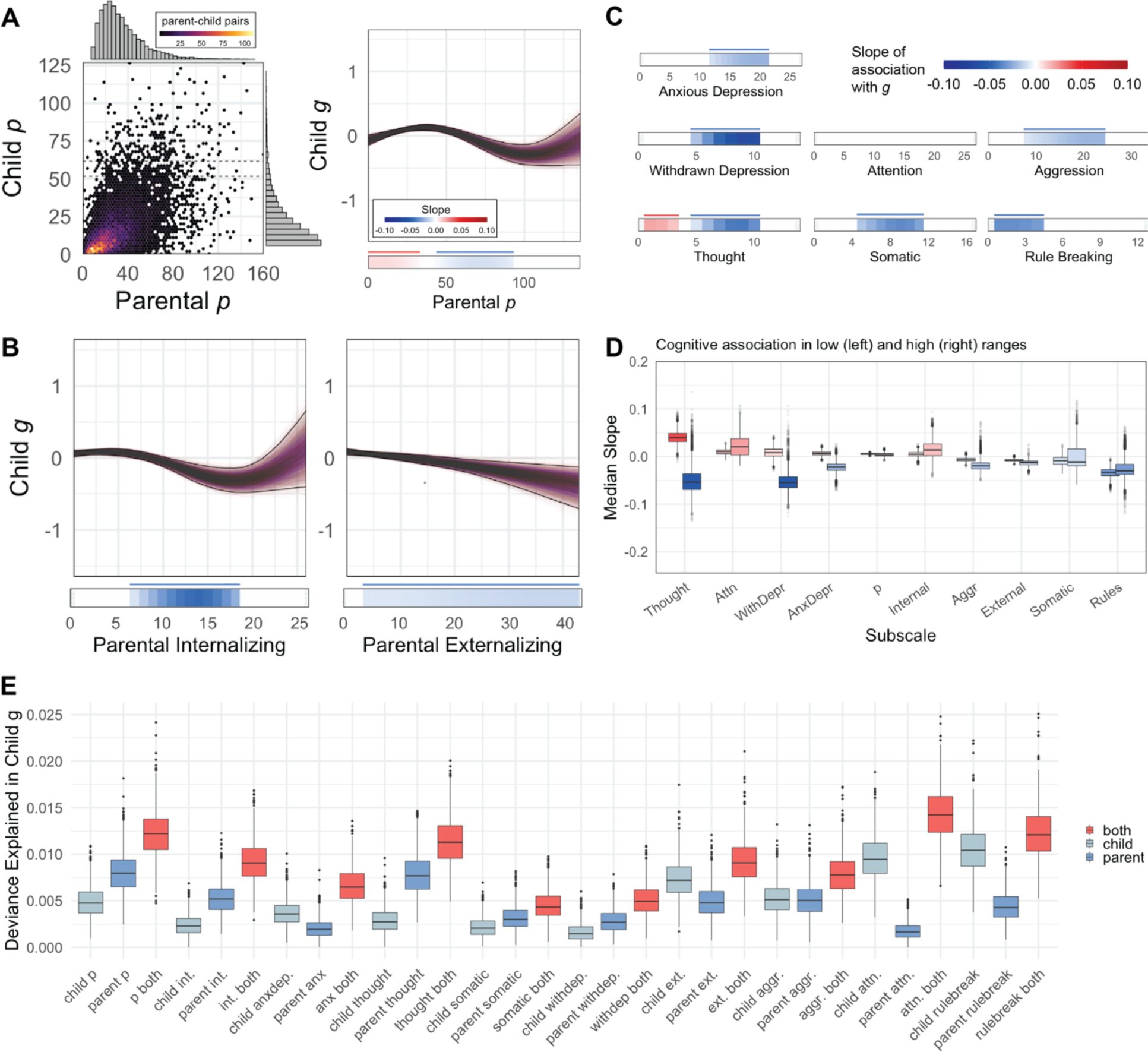
Child cognition is non-linearly associated with parental mental health. **A)** *Left*: Parental general psychopathology (*p*) is associated with *p* in children (*r* = 0.57, *p* < 0.001). *Right:* Penalized splines in 10,000 bootstrap resamples reveal a non-linear relationship between parental *p* and child *g*, such that lower quantities of reported symptoms in parents are *positively* associated with child cognition but higher quantities of symptoms are *negatively* associated with child cognition. **B)** Decomposition of parental general psychopathology into internalizing and externalizing reveals differences in what level of symptom severity cognitive deficits manifest at. A smooth, relatively consistent decline in cognition is observed with increasing externalizing symptoms, but internalizing symptoms are not associated with lower *g* at low levels of symptoms: negative associations between internalizing symptoms and *g* primarily manifest at moderate parental symptom levels. **C)** Further decomposition of parental psychopathology into constituent subscales reveals complex relationships with child cognition. Across subscales, the association between symptoms and *g* was stronger at certain levels of symptoms more than others, suggesting that a single slope does not fully capture the relationship between any one parental psychopathology dimension and child cognition. **D)** In the healthiest third of symptoms, most parental symptoms were positively associated with cognition. However, in the least healthy third of symptoms, parental symptoms were negatively associated with cognitive symptoms. **E)** Deviance in child *g* explained by child mental health, parental mental health, and both measures for each subscale. In all instances, covarying for both mental health measures explained more deviance in child cognition and resulted in better AIC (**Table S4**).

Importantly, we found that parental mental health explained unique variance in child cognition, above-beyond that explained by child mental health (**Figure 2E**). Further, for general psychopathology, internalizing, thought, somatic, and withdrawn depression, parental mental health explained more variance in child cognition than equivalent child measures of mental health. In all instances, inclusion of terms for both child and parental mental health resulted in better model performance (**Figure 2E; Table S4**). Together, these results suggest that the strength and direction of the association between parental mental health and child cognition is also systematically obscured by assuming a unitary fixed slope, and that parental mental health may constitute a critical dimension of heterogeneity in child mental health.

### Grades are a more generalizable indicator of child psychopathology than general cognition across sociodemographic heterogeneity

We next sought to disentangle the contributions of two other highly influential interactors in cognitive development: biological sex and socioeconomic status. In addition to constituting key sources of psychiatric heterogeneity and vulnerability, evidence suggests that sex and socioeconomic status interactions are systematically overlooked in assessing the generalizability of psychiatric research^71,72^. We therefore assessed the generalizability of relations between *g* and *p* across sex-assigned-at-birth (SAAB) and across groups of children above and below the federal poverty line. To contextualize differences in the association between *g* and *p* across these groups of children, we first confirmed concordance with prior literature in group differences in *p*. Boys on average exhibited more symptoms than girls at baseline (*t* = 7.17, *p* = 8.6×10^−13^) and two-year follow up visits (*t* = 5.68, *p* = 1.4×10^−8^), driven by greater externalizing symptoms. We recovered the expected negative association between child *p* and parental income at baseline (*t* = 5.32, *p* = 1.4×10^−7^) and two-year follow up visits (*t* = 4.50, *p* = 8.0×10^−6^, **Figure S8**).

Consistent group differences were also present in the slope of cognitive-symptom associations. Specifically, decline in cognitive scores was associated with fewer symptoms in boys than in girls. However, the negative relationship between cognition and symptoms was stronger and persisted over a greater span of symptoms in girls (**Figure 3A**). Qualitatively, co-existing positive and negative relationships between cognition and symptoms were more common in girls (**Figure S9**), suggesting that the degree to which general cognition linearly tracks with symptom burden might itself be sex-dependent. These results suggest that cognitive impairment in psychiatric illness might be detectable in boys with fewer symptoms, but that, when cognitive impairments occur in girls, the association between cognition and symptoms might be more profound and complex.

**Figure 3:**
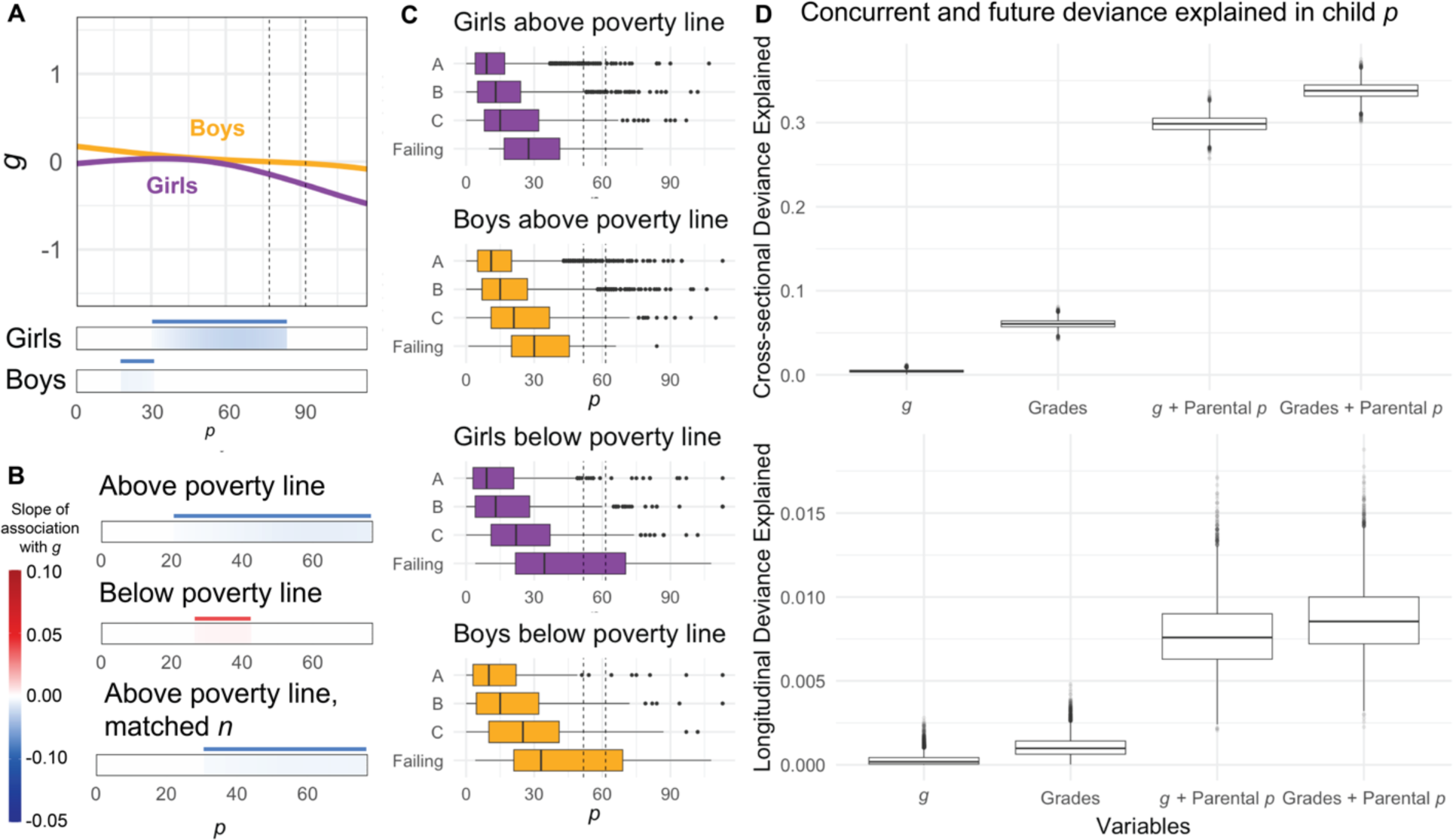
Grades predict psychopathology better than general cognition in children. **A)** The relationship between *g* and child *p* varies across SAAB, such that the slope of this relationship manifests more strongly and at greater symptom burdens in girls. Median model fits across bootstraps displayed for clarity. **B)** The relationship between *g* and *p* may depend on socioeconomic status: we were unable to recover any significant negative association between *g* and *p* in children below the federal poverty line (*n* = 617 children, 1,234 observations), and instead observed a *positive* association between *g* and *p*. Confidence intervals in this panel exclusively were evaluated at 90% (consistent slope direction in >90% of iterations) rather than 99% as for other analyses. **C)** Grades are an interpretable and generalizable proxy for healthy mental function in children. Grades were positively associated with cognition in both sexes above and below the poverty line. **D)** *Top:* Across bootstrap resamples, grades were more strongly associated with mental health cross-sectionally. *Bottom:* Grades were yielded slightly more deviance explained in timepoint 2 mental health than *g* in equivalent models. However, AIC-based model comparisons did not provide evidence for substantial contributions of *g* or Grades to timepoint 2 *p* predictions after accounting for timepoint 1 *p*.

Critically, despite inclusion of many children in poverty in the ABCD sample (*n* = 617 children, 1,234 observations), we were unable to recover any negative relationship between *g* and *p* in children below the federal poverty line across bootstrap iterations. Even when relaxing our criteria for statistical significance from 99% confidence to 90% confidence (**Figure 3B**), we only observed a *positive* association between *g* and *p* in children below the federal poverty line. In contrast, we were able to recover a negative relationship between *g* and *p* both in the complete sample of children above the poverty line and in bootstrap iterations of children above the poverty line constrained to be the same number of children sampled below the poverty line (**Figure 3B**). Strikingly, when repeating the same analyses over all CBCL and ASR subscales, no consistent negative slopes between cognition and symptoms were found in children below the poverty line. This lack of generalization was apparent for every mental health subscale in both children (**Figure S9**) and parents (**Figure S10**), where only positive relationships between cognition and symptom burden were discovered (confidence interval ≥ 99% in 13/18 subscales). These results suggest that the assumed canonical relationship between cognition and psychopathology (impaired cognition with more symptoms) may not generalize to children living in poverty. An alternative possibility, as discussed in the proceeding section, is that we were unable to recover a negative relationship between *g* and symptoms in children below the poverty line due to sampling bias.

Next, we addressed psychopathology in impaired daily functions. If impaired daily functions are a “common denominator” for how cognitive impairment gives rise to mental health deficits, it follows that impaired daily functions might serve as a more generalizable measurement for mental health deficits across diverse groups of children. After ensuring that grades exhibited the expected relationship to *g* (higher *g* = higher grades, *p* <0.001, **Figure S11**), we did observe that scholastic grades provided a proxy for psychiatric well-being in children. Unlike the relationship between general cognition and symptoms, the relationship between grades and symptoms clearly generalized across sociodemographic categories including boys and girls both above and below the federal poverty line (**Figure 3C**, all *p*s < 0.001).

Finally, we tested whether grades were more associated (variance in *p* explained within-timepoint) and predictive (variance explained in timepoint 2 mental health after controlling for timepoint 1 mental health) of mental health than the *g* factor. For these analyses, we considered psychopathology scores as an outcome variable instead of an independent variable. Comparisons between within-timepoint models revealed that grades better-explained variance in psychopathology than did *g* factor scores (**Figure 3D**, top, **Table S5**). Although AIC model comparisons between future-timepoint models did not suggest that timepoint 1 *g* and grades explain timepoint 2 *p* above-and-beyond timepoint 1 *p* (**Table S5**), deviance explained in timepoint 2 *p* was consistently higher in models including grades relative to models including *g* (**Figure 3D**, bottom). Together, these results suggest that compared to *g*, grades may provide a more generalizable and accurate indicator of mental health deficits in children.

## Discussion

### Positive and negative associations between cognition and psychopathology co-exist

We found that cognition was both positively and negatively associated with psychiatric symptoms in children, depending on the class and level of symptom burden. By relaxing the assumption that cognition and psychopathology have a fixed linear correspondence across diverse presentations of psychopathology, these findings resolve conflicting prior reports of negative^34–37^ or positive^39–43^ associations between cognition and psychopathology. Importantly, our results do not invalidate either set of findings: we uncovered evidence for *both* positive and negative associations between symptoms and cognition. Rather, our results suggest that divergent findings may stem from an interaction between variable clinical enrichment across studies and use of models assuming a one-size-fits-all slope, given that cognitive associations depends on the level of symptom burden. Further, because these cognitive-symptom relationships can be opposed at different levels of symptoms, aggregating a linear slope across the entire spectrum of symptoms negates the derived strength of the association between cognition and symptoms. An assumption latent in linear modeling is that the relationship between the independent and dependent variable is constant across the entire range of the independent variable. Here, we provide evidence that this assumption is violated for the relationship between cognition and psychopathology.

It follows that in studies without clinical enrichment, it is entirely feasible to recover a positive relationship between *g* and mental health symptoms with linear models, as symptoms are positively associated with general cognition at low levels of symptoms. Notably, clinically enriched studies consistently recover negative associations between cognition and symptoms^35,37,38^, whereas positive associations between cognition and symptoms have been primarily observed in studies enriched for high-functioning participants^39,42,43^. However, a central goal of studying cognition in humans is to improve understanding and treatment of cognition specifically in mental illnesses. Because the relationship between cognition and symptoms is robustly negative at clinical symptom burdens but is greatly weakened or even inverted at low symptom burdens, our results make a clear case for the necessity of clinical enrichment in studies aiming to study cognition in mental health.

### Parental psychopathology as a dimension of clinical heterogeneity in children

Here, we present evidence that parental psychopathology might constitute a key dimension of cognitive heterogeneity in children. Parental mental health exhibited a complex relationship with child cognition, and for general and internalizing symptoms, was more strongly related to child cognition than child mental health itself. However, this finding should be interpreted with care. Internalizing symptoms are difficult to accurately report, and parental difficulties in reporting their children’s internalizing symptoms are well-documented^73^. Although child-reported psychopathology is not necessarily more accurate than parental report of child psychopathology^59^, parents can certainly miss or misunderstand symptoms experienced by their children^74^. It is possible that in the current study, a higher degree of deviance explained in child cognition by parental internalizing symptoms might be more attributable to measurement accuracy than mental health mechanisms. Regardless, parental mental health was found to be a strong correlate of concurrent child mental health, and predictive of future child cognition above-and-beyond concurrent measurement of child mental health. Consequently, in addition to aiming for clinical enrichment among children, future studies incorporating parental mental health may lead to more accurate characterizations of psychiatric cognitive impairments in children. Although we cannot causally demonstrate that strong parental mental health elevates cognitive development in children, our results serve as observational evidence that parental mental health interventions may benefit cognitive development in children^75^. In instances where parental mental health might be a central factor in a child’s mental health, a precision intervention *for the child* might be to address parental mental health rather than solely treating the child’s symptoms^76–78^.

### Limitations in generalizability demonstrate the need for precision psychiatry

Psychiatric research has increasingly recognized the importance of individual differences in psychopathology in patients above and beyond their reported symptoms^7,10^. Because treatment efficacy and endorsed symptoms vary dramatically within diagnostic categories^3^, it is critical to understand the boundaries of generalizability to account for individual variability beyond such boundaries. As an initial step, coarse, demographic measures of individual differences are key considerations that serve as context for finer-grained heterogeneity^79^. Here, we demonstrate that the relationship between cognition and psychopathology does not generalize across key demographic variables and highlight the need for considering cognitive heterogeneity *at least* at the level of SAAB and family income. In fact, we were only able to recover a *positive* relationship between *g* and psychopathological burden for children below the poverty line: the opposite of what has been predominantly reported^35^. This discrepancy was more evident at the level of CBCL and ASR subscales. For children in poverty no negative relationships between symptoms and cognition were found across 21 parent and child mental health measures. Unfortunately, this limited generalizability of the association between *g* and *p* is consistent with limited generalizability of treatment efficacy to individuals with low socioeconomic status^80,81^. Ultimately, a goal of precision psychiatry is to extend diagnostic understanding and treatment efficacy beyond these limits of generalizability. In addition to incorporating clinically enriched samples and parental mental health measurements, recruiting, retaining, and working with socioeconomically diverse participants may be necessary to determining the generalizability of cognitive-symptom associations.

### Grades provide a more generalizable and accurate measurement of preclinical psychiatric risk

Finally, in lieu of full clinical evaluation and delineation, grades may constitute an actionable, generalizable, and accurate marker of psychiatric health children. These findings cohere with reports of life satisfaction being more directly tied to grades than direct tests of academic competencies in German schoolchildren^5^. Although the exact link between grades and psychopathology is likely complex, one reason for the relatively strong association between grades and symptoms may be their proximity to daily function and performance in children. Mental health is often defined partly on the basis of performance (or failure to perform) in the environment of the individual^82,83^. For children, grades reflect their performance in their primary non-home environment, which in turn is coupled to peer, familial, societal, and self-worth^5,33,82^. Whether or not a child’s self-worth *should* be tightly coupled to the grades, our study provides evidence that they *are* coupled to mental health across major demographic stratifications.

### Limitations

Our study is also subject to several limitations. First, as we have noted, our research is observational. Child psychopathology cannot be experimentally induced, but interventions aimed at alleviating cognitive impairments in children may elucidate causality. Second, although non-causal, additional and more frequent measurement timepoints may afford greater clarity in temporal precedence. Third, every study is subject to potential sampling biases. Although ABCD is nearly unprecedented in socioeconomic diversity among neurocognitive studies, it is not possible to fully rule out that children in certain environments were disproportionately excluded from the present study. It is possible that undetected sampling biases could have interacted with our results. For example, we only detected a positive slope between *g* and *p* for children below the federal poverty line: it’s possible that participation in ABCD is particularly infeasible for families below the poverty line with children suffering from both impaired cognition *and* clinical symptom burdens. Such an unseen but systematic participant-selection pressure could artificially inflate a positive association between symptoms and cognition, as we have observed, in children currently below the poverty line. To this point, we identified lower-than-expected representation of Asian American children in our sample both before and after exclusions were applied. Future datasets enriched for Asian Americans^84^ and extant datasets enriched for Asian individuals^85–87^ may aid in determining the broad generalizability of psychiatric relationships. However, the ABCD study was painstakingly constructed to minimize sampling bias^51^, and to our knowledge, is currently unmatched in its diversity among similar samples. In addition to these efforts, more cohorts^84–86^, resources^61,88^, and researchers^89–91^ will be needed to fully determine the generalizability of neuropsychiatry findings.

### Conclusion

Our results suggest that the relationship between cognition and psychopathology is complex: full delineation of this complexity will likely require samples with substantial clinical enrichment, parental mental health, and diverse socioeconomic representation. Here we demonstrate that class and magnitude of symptoms, parental mental health, SAAB, and poverty constitute at least five dependencies in the relationship between cognition and psychopathology. In light of this complexity, grades may serve as a more efficacious proxy of psychiatric health in children.

## Data Availability

All data is from the ABCD study. All analysis code is in the public domain.

## Acknowledgements

A.P. was funded by the Stanford School of Medicine Dean’s Fellowship and through a Gustavus and Louise Pfeiffer Research Foundation award to L.M.W. The funders of the ABCD study had no role in data analysis, data interpretation, or writing of the report. Data used in the preparation of this article were obtained from the Adolescent Brain Cognitive Development® (ABCD) Study (https://abcdstudy.org), held in the NIMH Data Archive (NDA). This research was supported by the NIH (ABCD Study; U01DA041174). The ABCD Study® is supported by the National Institute of Health and Additional Federal Partners under award numbers: U01DA041048, U01DA050989, U01DA051016, U01DA041022, U01DA051018, U01DA051037, U01DA050987, U01DA041174, U01DA041106, U01DA041117, U01DA041028, U01DA041134, U01DA050988, U01DA051039, U01DA041156, U01DA041025, U01DA041120, U01DA051038, U01DA041148, U01DA041093, U01DA041089, U24DA041123, U24DA041147. A full list of supporters is available at https://abcdstudy.org/federal-partners.html. A listing of participating sites and a complete listing of the study investigators can be found at https://abcdstudy.org/consortiummembers/. ABCD consortium investigators designed and implemented the study and/or provided data but did not necessarily participate in the analysis or writing of this report. This manuscript reflects the views of the authors and may not reflect the opinions or views of the NIH or ABCD consortium investigators. We are extremely grateful to the study participants for their time and participation.

---

**Figure S1:**
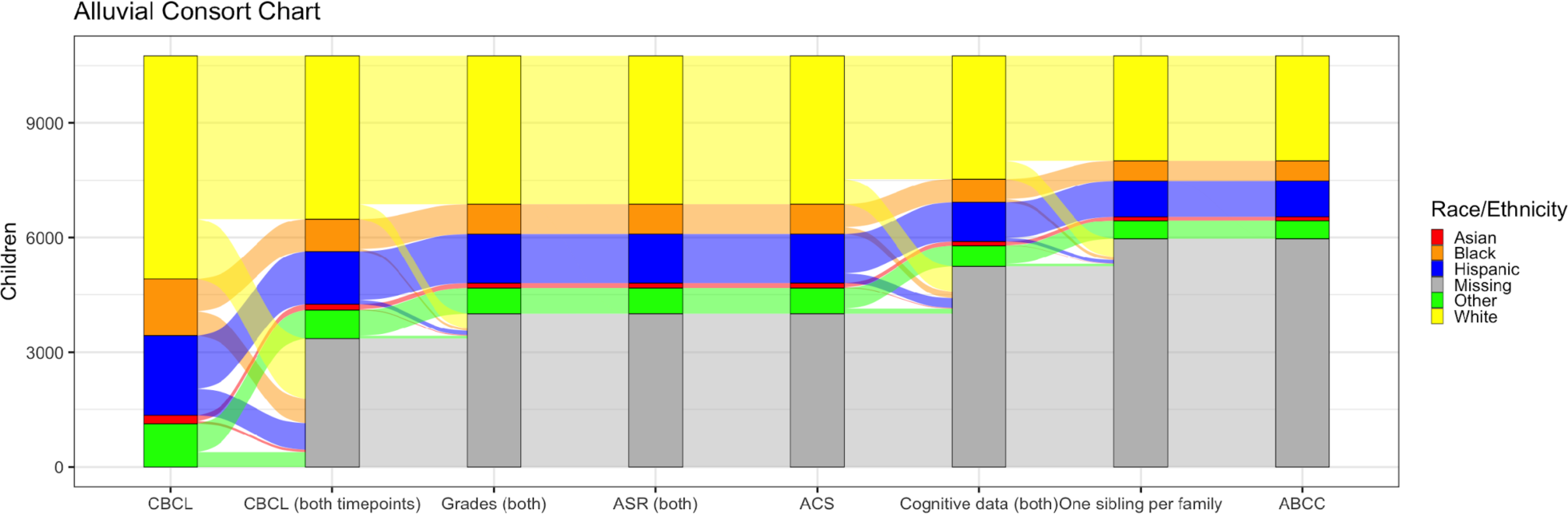
Study exclusion criteria. Of 11,827 participants (far left) with child behavioral checklist (CBCL) data at baseline assessment, 3,764 were excluded for not having CBCL data at both timepoints. Of the remaining 8,063, 715 participants were excluded for not having scholastic (Grades) data at both timepoints. Of the remaining 7,348, an additional participant was excluded for not having adult self-report (ASR) data at both timepoints, and an additional participant was excluded for not having family information present (ACS). Of the remaining 7,346 participants, 1,351 were excluded for not having recorded cognitive measures at both timepoints. Of the remaining 5,994 children, 780 were removed for having siblings within the study. Of the remaining 5,214 participants, 44 were excluded for inconsistent reporting of participant site.

**Figure S2:**
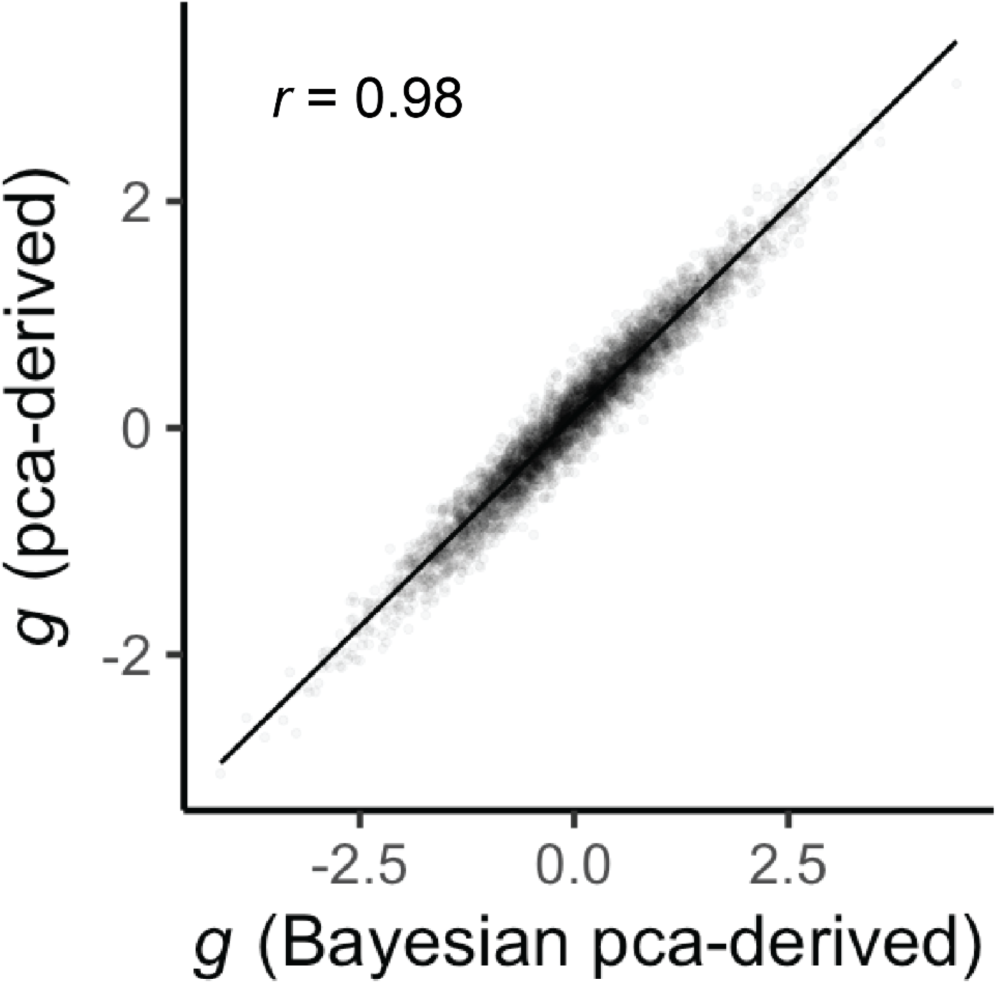
Similarity of *g* factorized from principal component analyses versus Bayesian principal component analysis. For general cognition, we modeled our approach after Thompson et al., 2019^55^. We demonstrate that the first principal component of these cognitive data yields a functionally equivalent measurement to the values derived from Bayesian principal component analysis in Thompson et al. (*r =* 0.98, *p* < 0.001).

**Figure S3:**
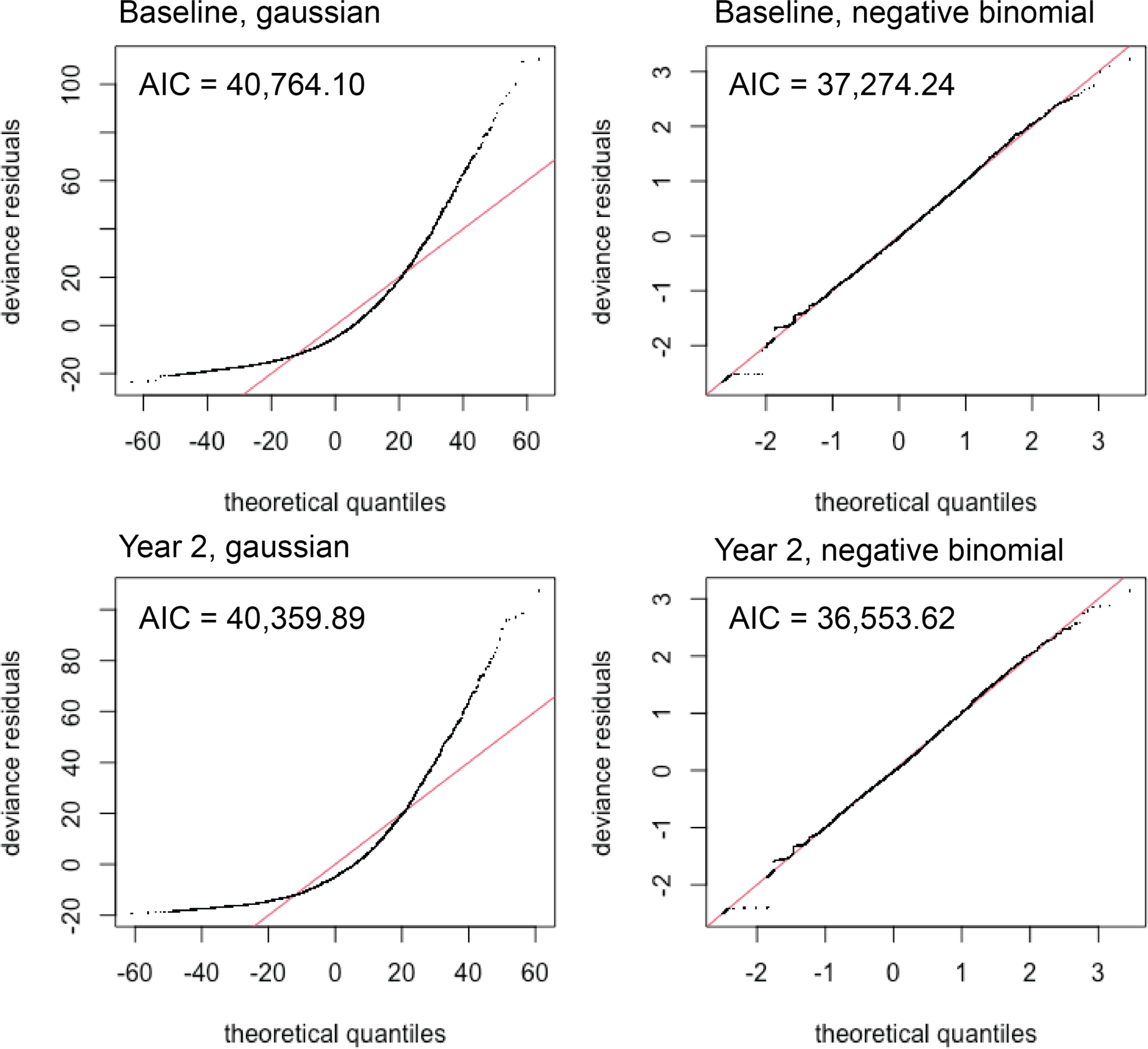
Negative binomial link functions provide a better fit to the relationship between *g* and *p*. Q-Q plots and AIC and BIC for baseline and two-year follow up data modeled as *p*∼s(*g*) with gaussian versus negative binomial link functions. AIC and BIC were consistently lower for negative binomial link functions.

**Figure S4:**
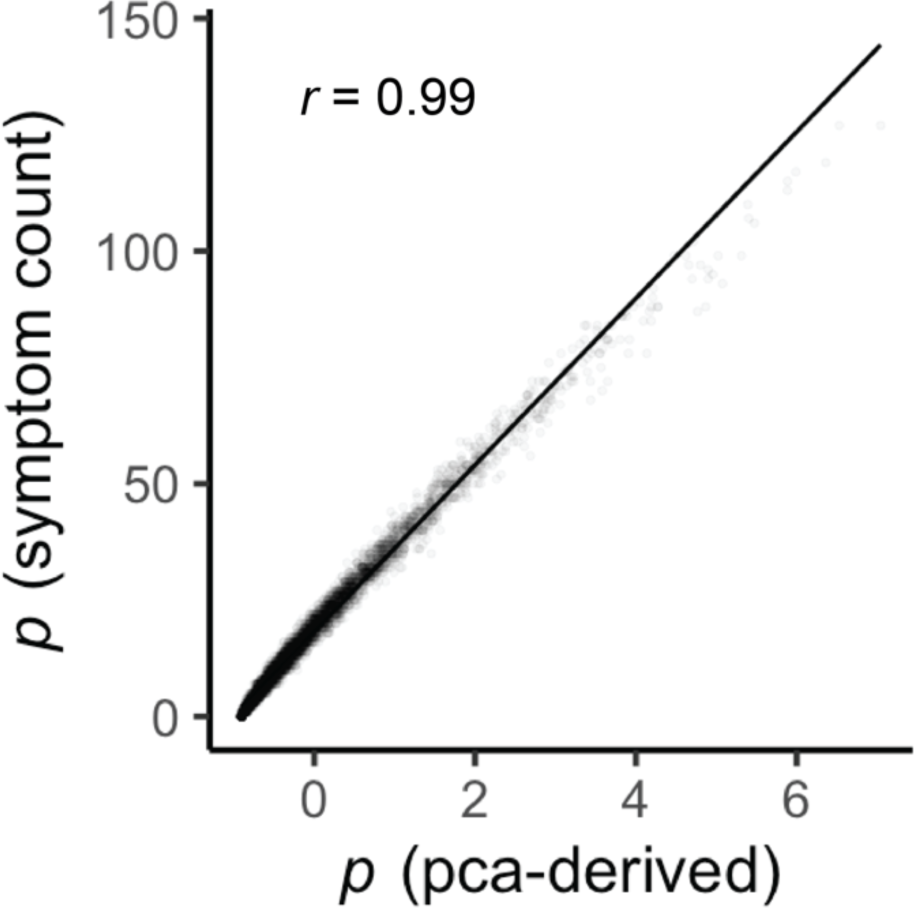
Similarity of symptom counts and formal p-factorization. We replicated an approach to deriving a formal general psychopathology factor (*p)* in accordance with Michelini et al., 2019^60^. Raw count of endorsed symptoms was *r* = 0.99 correlated with principal-component-derived *p* factors, suggesting that the raw symptom count is functionally equivalent to that derived from principal component analysis.

**Table S1:** Temporal precedence in cognition and psychopathology. Using the longitudinal timepoints available in ABCD, we conducted a cross-lagged panel analysis to test for temporal precedence between *g* and psychopathology variables. Full-model results recapitulated prior findings^21,62,63^: psychopathology and cognition are likely both causal of each other, but our evidence suggests that timepoint one psychopathology variable explains more variance in timepoint two cognition variables than vice-versa (cognitive variables in timepoint 1 explaining variance in timepoint 2 psychopathology).

| Timepoint 1 -> Timepoint 2 | Cross-lagged effects (z) | *p* |
| --- | --- | --- |
| *g* -> *p* | 0.68 | 0.500 |
| *g* -> Internalizing | 2.81 | 0.005 |
| *g* -> Externalizing | -1.39 | 0.166 |
| *p* -> *g* | -2.10 | 0.035 |
| Internalizing -> *g* | 0.24 | 0.814 |
| Externalizing -> *g* | -3.07 | 0.002 |

**Table S2:** AIC from linear and nonlinear comparison models for all CBCL subscales. Lower AIC in otherwise equivalent nonlinear models (i.e., *g* ∼ *s(p)* + *s*(Age) vs. *g* ∼ *p* + *s*(Age)) suggests that the relationship between general cognition and symptoms is better-explained by a nonlinear association for *p*, internalizing, anxious depression, withdrawn depression, thought, social, and somatic subscales. Conversely, comparison of AIC between equivalent models suggested that linear fits adequately capture the association between *g* and externalizing, attention, aggression, and rule-breaking symptoms.

| Scale | AIC (linear) | AIC (nonlinear) |
| --- | --- | --- |
| $g \sim s(p)$ | 26242.94 | 26237.54 |
| $g \sim s(\text{Internalizing})$ | 26265.45 | 26243.8 |
| $g \sim s(\text{Externalizing})$ | 26214.1 | 26214.29 |
| $g \sim s(\text{Anxious Depression})$ | 26245.84 | 26224.38 |
| $g \sim s(\text{Withdrawn Depression})$ | 26268.38 | 26261.42 |
| $g \sim s(\text{Thought})$ | 26264.4 | 26237.91 |
| $g \sim s(\text{Social})$ | 26168.58 | 26167.41 |
| $g \sim s(\text{Attention})$ | 26192.55 | 26192.55 |
| $g \sim s(\text{Somatic})$ | 26260.71 | 26254.34 |
| $g \sim s(\text{Aggression})$ | 26234.42 | 26234.75 |
| $g \sim s(\text{Rule Breaking})$ | 26178.49 | 26178.49 |

**Figure S5:**
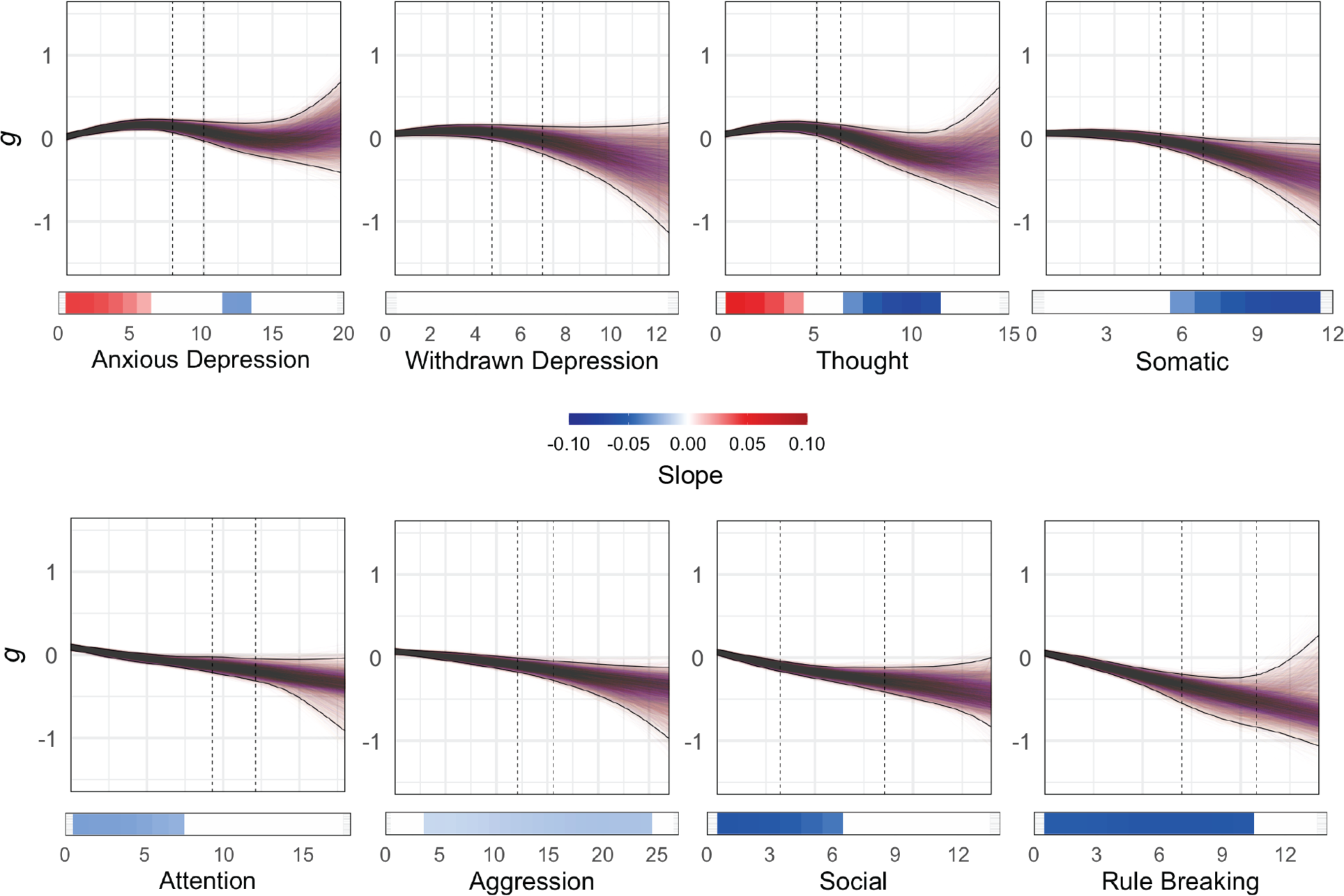
Bootstrap iterations of the relationship between *g* and psychopathology in all subscales in children. Derivatives (colored bars) are identical to those presented in the main text. Bootstrap iterations revealed a broad procession from *positive* associations with *g* at low levels of symptoms to *negative* associations with *g* at high levels of symptoms, particularly within internalizing classes of symptoms.

**S6:**
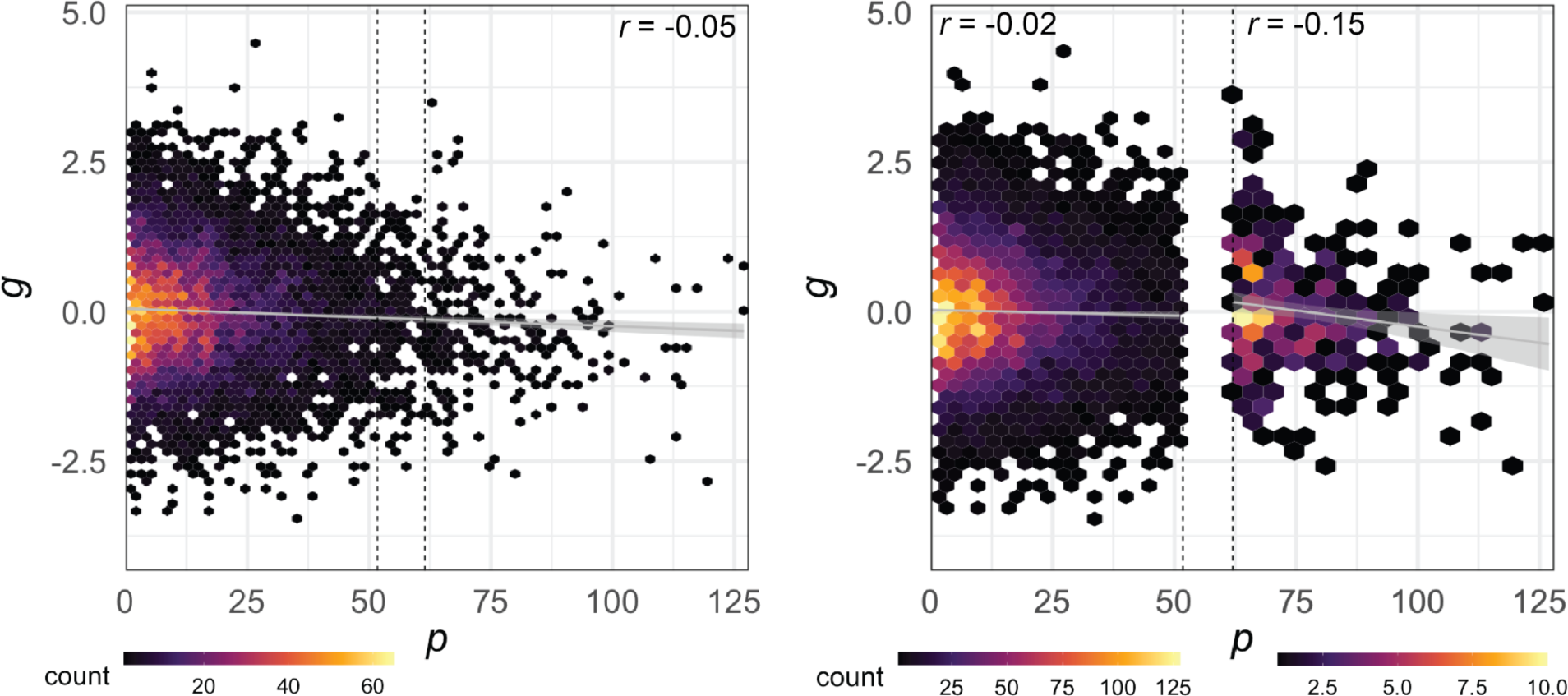
Linear relationship between general cognition (*g*) and psychopathology (*p*) with and without clinical enrichment. As a proof of concept, we derived *g ∼ p* correlations specifically above and below clinical thresholds. The effect size estimated for general cognition and psychopathology is 8 times larger in the clinical subset than that observed across the entire sample (*r*/*r*^2^ = −0.052/0.003 vs. *r*/*r*^2^ = −0.149/0.022), and 44 times larger than an effect size derived purely within the subclinical range of the sample (*r*/*r*^2^ = −0.023/0.0005 vs. *r*/*r*^2^ = −0.149/0.022).

**Table S3:** AIC from linear and nonlinear comparison models for all ASR subscales. Lower AIC in otherwise equivalent nonlinear models (i.e., child *g* ∼ *s(parental p)* + *s*(child age) vs. *g* ∼ *p* + *s*(child age)) suggests that the relationship between child general cognition and parental symptoms is better-explained by a nonlinear association for *p*, internalizing, anxious depression, withdrawn depression, thought, social, somatic, aggression, and attention subscales. Conversely, comparison of AIC between equivalent models suggested that linear fits adequately capture the association between child *g* and parental externalizing and rule-breaking symptoms.

| Scale | AIC (linear) | AIC (nonlinear) |
| --- | --- | --- |
| Child $g \sim s(\text{Parental } p)$ | 26265.3 | 26206.17 |
| Child $g \sim s(\text{Parental Internalizing})$ | 26245.8 | 26223.19 |
| Child $g \sim s(\text{Parental Externalizing})$ | 26236.54 | 26236.85 |
| Child $g \sim s(\text{Parental Anxious Depression})$ | 26266.56 | 26256.62 |
| Child $g \sim s(\text{Parental Withdrawn Depression})$ | 26259.85 | 26249.96 |
| Child $g \sim s(\text{Parental Thought})$ | 26255.37 | 26210.82 |
| Child $g \sim s(\text{Parental Attention})$ | 26266.71 | 26261.24 |
| Child $g \sim s(\text{Parental Somatic})$ | 26245.29 | 26241.9 |
| Child $g \sim s(\text{Parental Aggression})$ | 26240.35 | 26238.22 |
| Child $g \sim s(\text{Parental Rule Breaking})$ | 26236.96 | 26236.96 |

**Figure S7:**
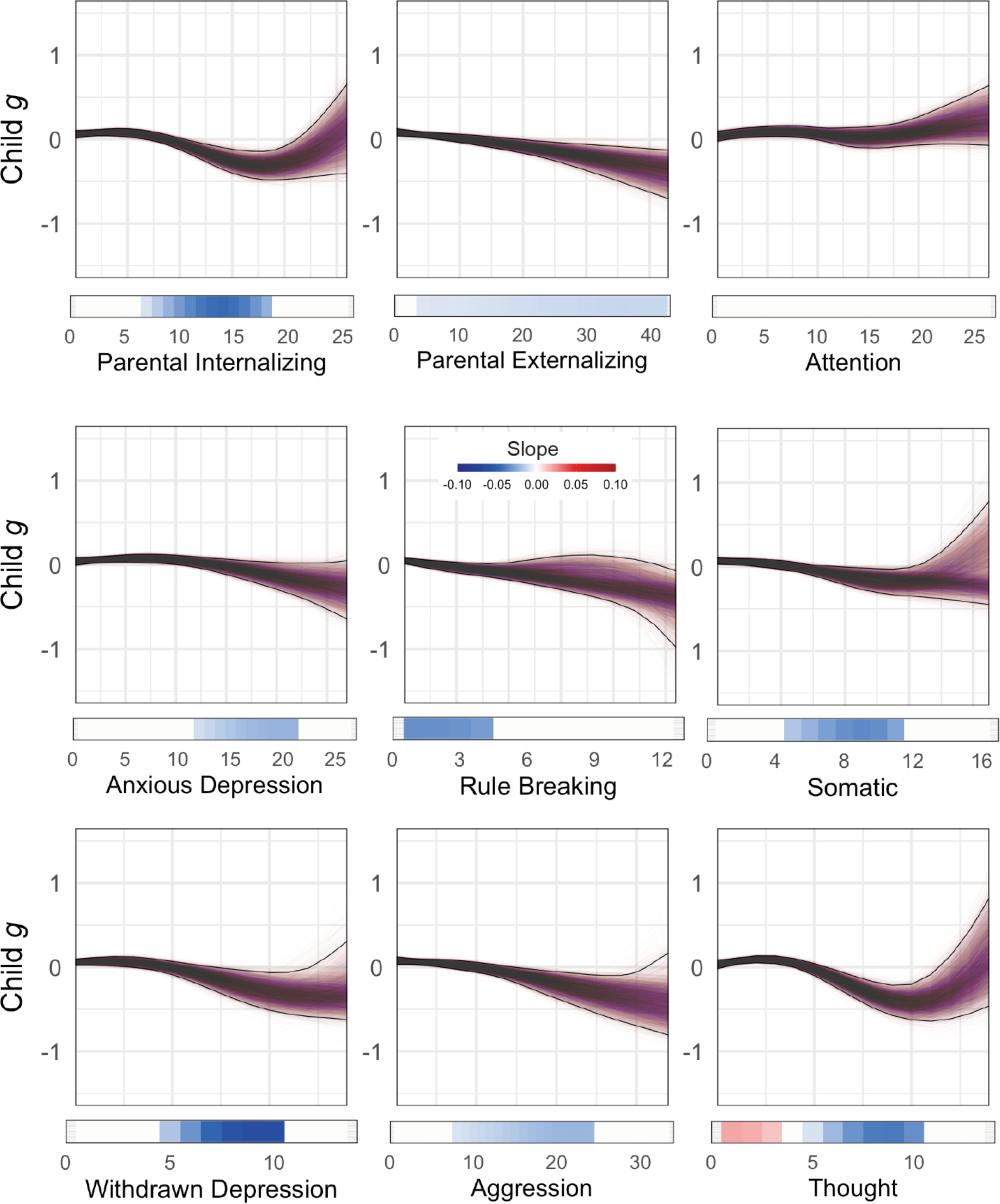
10,000 bootstrap iterations of psychopathology subscales in parents. Derivatives (colored bars) are identical to those presented in the main text. Bootstrap iterations consistently reveal symptom burden-dependent relationships to child cognitive scores, such that low levels of parental symptoms were less associated with cognition than higher levels of parental symptoms.

**Table S4:** AIC from models explaining child *g* comprised of terms for child symptoms, parent symptoms, and both sets of symptoms. Across all subscales, inclusion of parent and child covariates for mental health improved AIC. Age was excluded as a covariate from these models to measure deviance in *g* explained purely by mental health (**Figure 2E**).

| **Scale** | **AIC (child)** | **AIC (parent)** | **AIC (both)** |
| --- | --- | --- | --- |
| $g \sim s(p)$ | 26979.39 | 26949.93 | 26915.06 |
| $g \sim s(\text{Internalizing})$ | 27003.61 | 26974.94 | 26945.91 |
| $g \sim s(\text{Externalizing})$ | 26951.80 | 26976.68 | 26941.36 |
| $g \sim s(\text{Anxious Depression})$ | 26990.82 | 27005.90 | 26970.60 |
| $g \sim s(\text{Withdrawn Depression})$ | 27010.72 | 26998.83 | 26986.24 |
| $g \sim s(\text{Thought})$ | 27000.45 | 26951.47 | 26928.18 |
| $g \sim s(\text{Attention})$ | 26930.79 | 27009.15 | 26895.84 |
| $g \sim s(\text{Somatic})$ | 27004.01 | 26995.36 | 26989.31 |
| $g \sim s(\text{Aggression})$ | 26973.25 | 26977.47 | 26957.70 |
| $g \sim s(\text{Rule Breaking})$ | 26921.80 | 26980.15 | 26909.32 |

**Figure S8:**
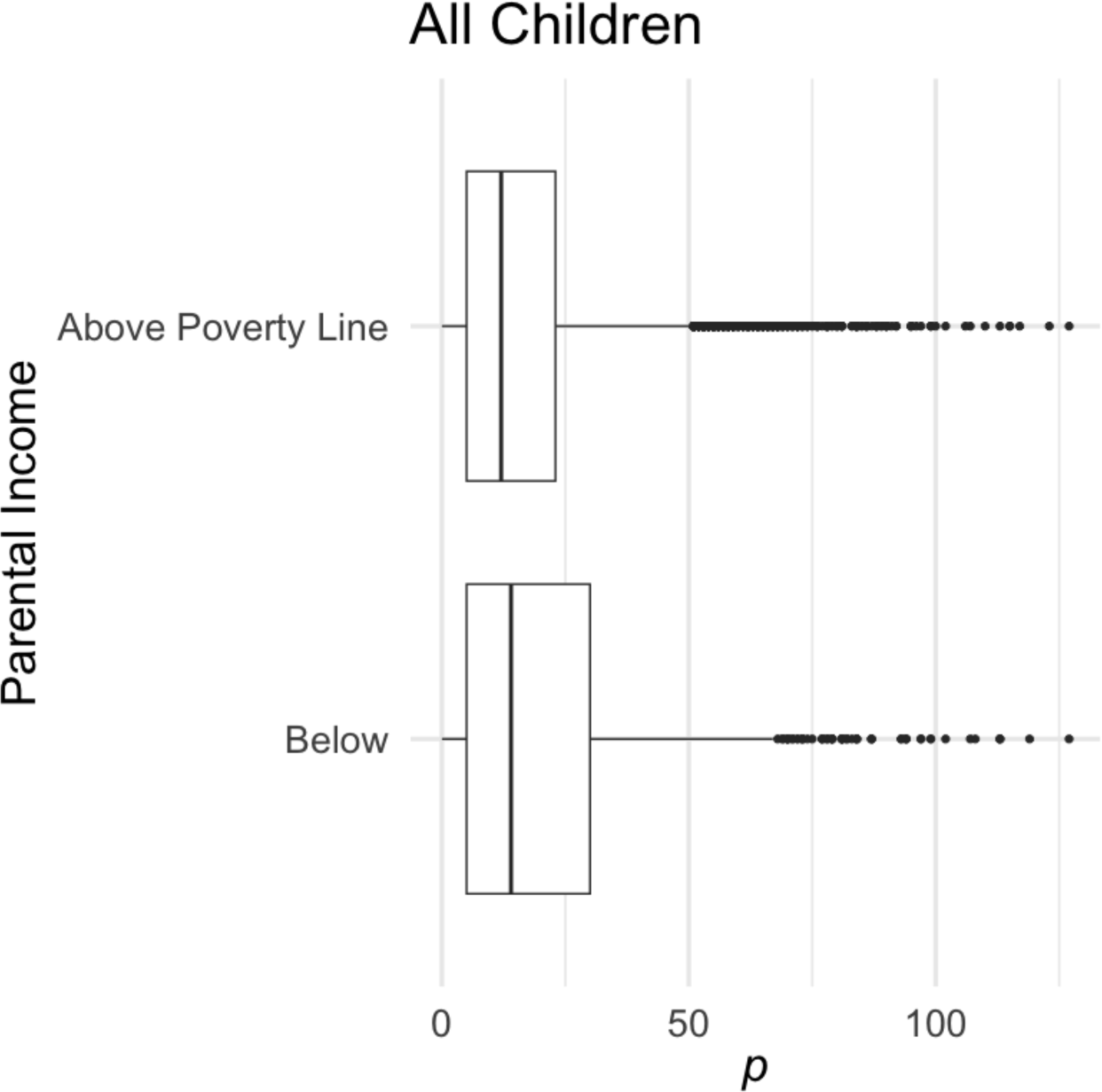
The relationship between general psychopathology (*p*) and poverty in children. Children from households falling below the federal poverty line exhibited more symptoms than children from households above the federal poverty line. This was true at baseline assessment (median symptoms = 12 vs. 16, *t* = 5.32, *p* = 1.4×10^−7^) and timepoint 2 (median symptoms = 11 vs. 13, *t* = 4.50, *p* = 8.0 x 10^-6^).

**Figure S9:**
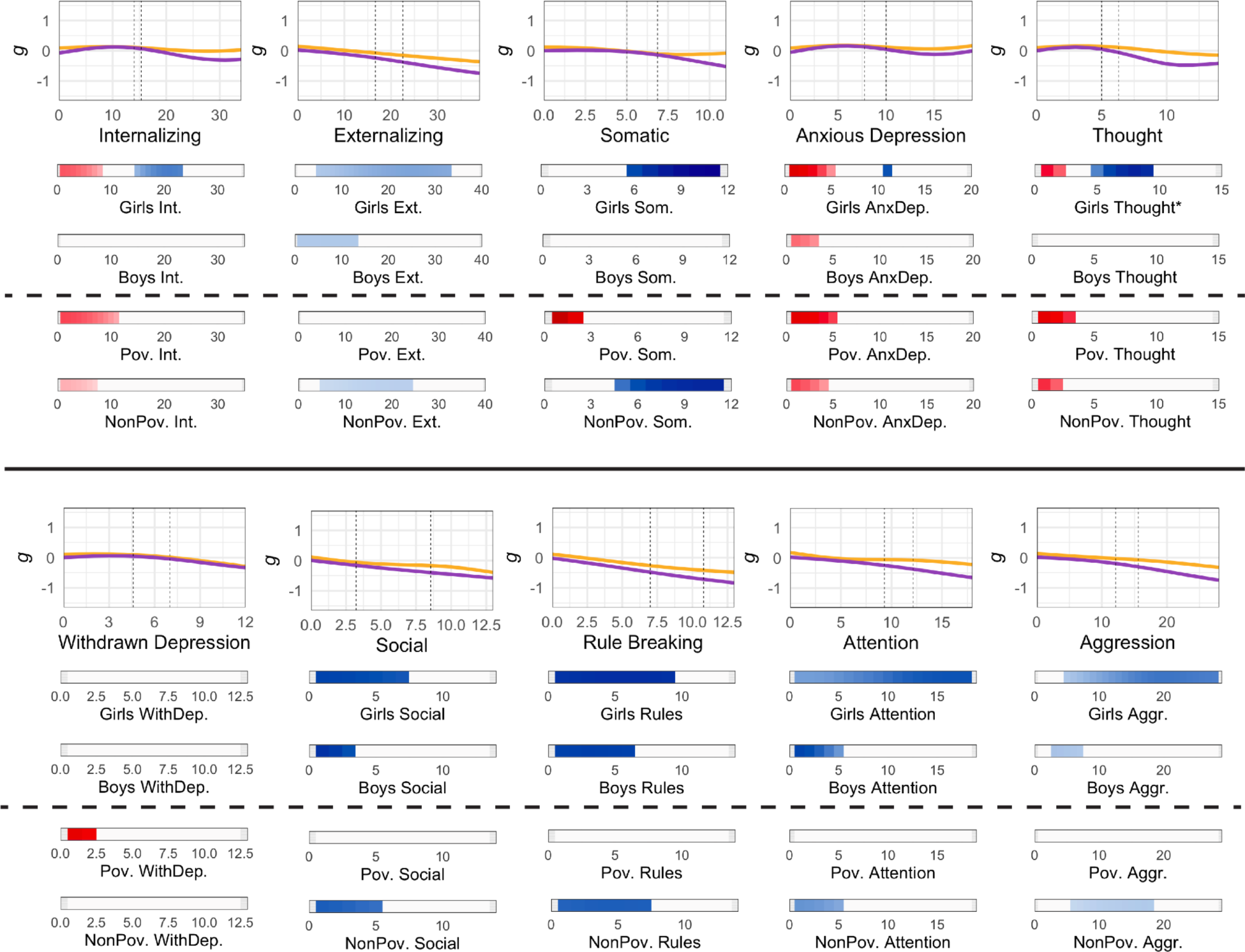
Bootstrap iterations of CBCL subscales across Girls/Boys (SAAB) and above/below the poverty line. Fits (orange/purple lines) and derived slopes meeting 99% confidence interval criteria (consistently positive or negative across > 9,900 bootstraps) are displayed for each subscale. Slopes above dashed lines correspond to SAAB, and slopes depicted beneath dashed lines correspond to significant slopes in children below and above the approximate federal poverty line. As in **Figure 3**, median fits are plotted for clarity. Asterisk (*) for Girls Thought indicates an expanded color mapping with −.15 (blue) and .15 (red) as the minimum and maximum slope, as the slopes for Girls Thought exceeded the maximal values of −.1 (blue) and .1 (red) used to denote slope throughout.

**Figure S10:**
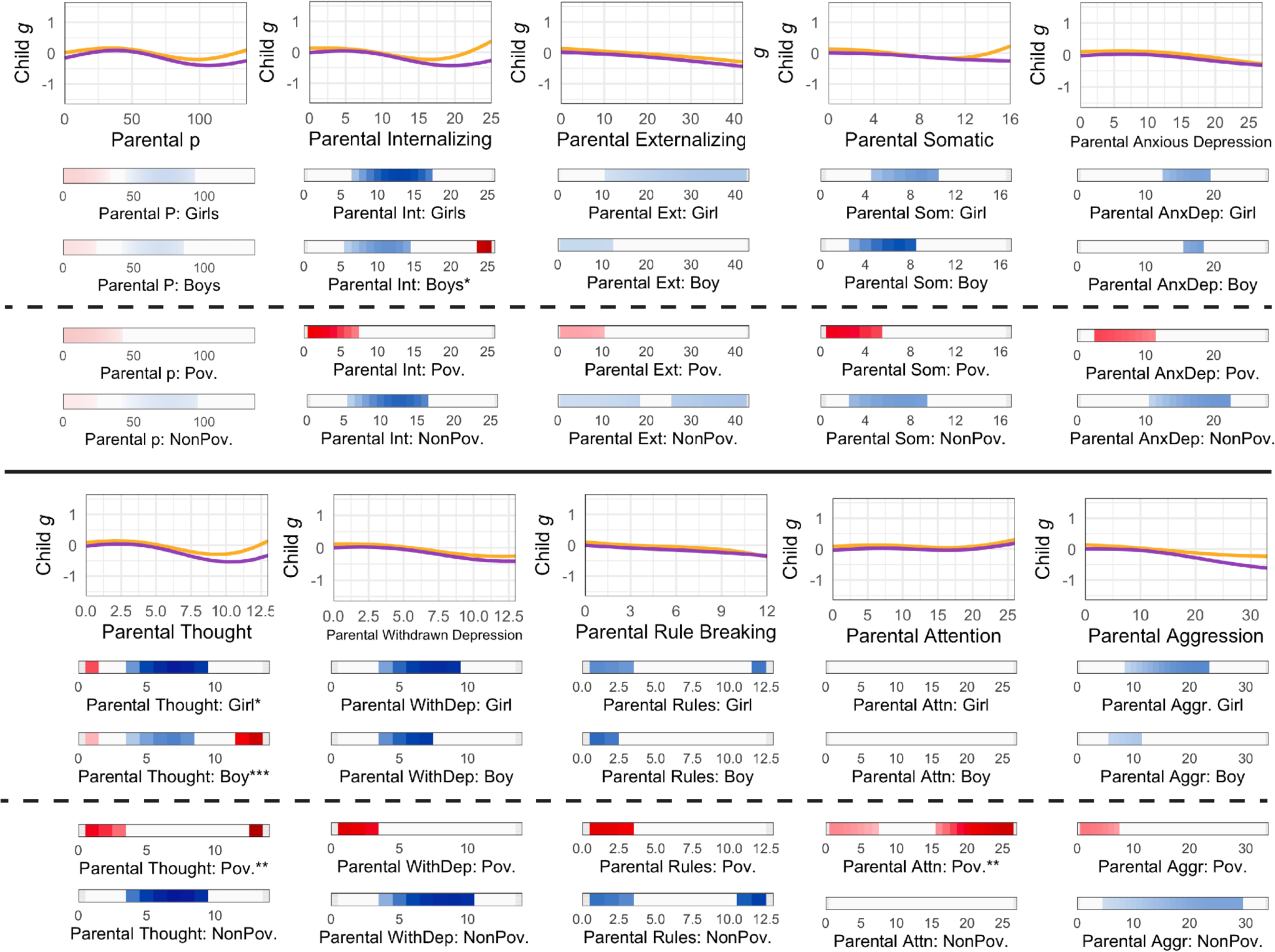
Bootstrap iterations of ASR subscales across Girls/Boys (SAAB) and above/below the poverty line. Fits (orange/purple lines) and derived slopes meeting 99% confidence interval criteria (consistently positive or negative across > 9,900 bootstraps) are displayed for each subscale of parental mental health. Slopes above dashed lines correspond to SAAB, and slopes depicted beneath dashed lines correspond to significant slopes in children below and above the federal poverty line. As in **Figure 3**, median fits are plotted for clarity. Asterisk (*) for Parental Internalizing: Boy and Parental Thought: Girl indicates an expanded color mapping with −.15 (blue) and .15 (red) as the minimum and maximum. Two asterisks (**) indicate that a greater expansion of color mapping was needed to capture sociodemographic-specific slopes (-.2 and .2, Parental Thought: Poverty and Parental Attention: Poverty), and three asterisks indicate that an even greater color range (-.3 and .3) was needed to capture slopes for child cognition ∼ parental thought problems in boys.

**Figure S11:**
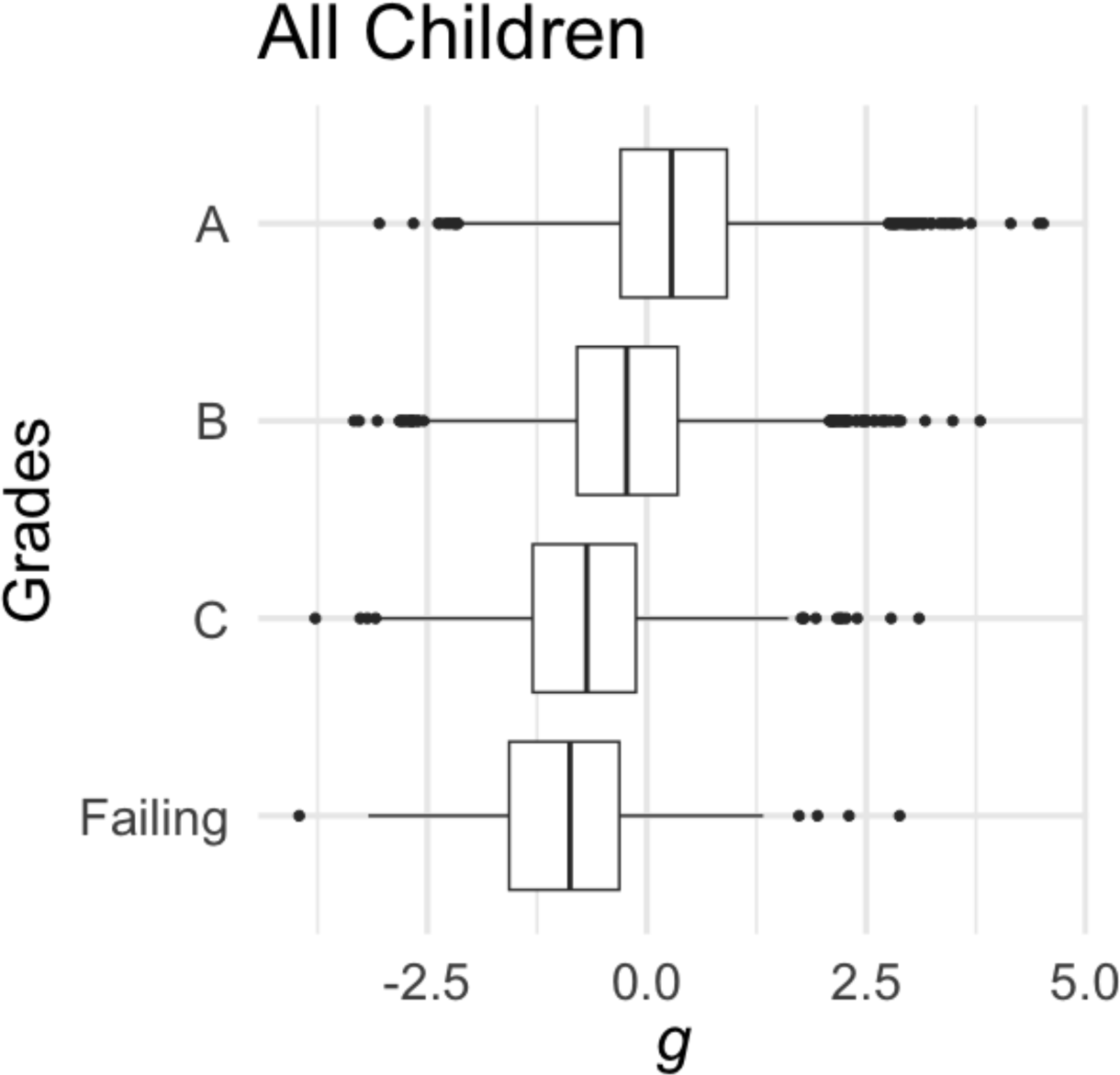
The relationship between general cognition (*g*) and scholastic grades in children. Grades were associated with cognition before and after covarying for age, such that higher *g* was associated with better grades (*ps* < 0.001). Average *g* for A’s, B’s, C’s, and failing at baseline = 0.153, −0.370, −0.878, and −1.248, respectively. Average *g* for A’s, B’s, C’s, and failing at timepoint 2 = 0.484, −0.03, −0.446, and −0.522.

**Table S5:** AIC from models explaining child *g* comprised of terms for child symptoms, parent symptoms, and both sets of symptoms. Model comparison of *p*-predicting covariates suggest that Grades better-explain individual variability in the *p* factor than does the *g* factor. Grades outperform *g* both with and without covarying for parental *p* factor scores. To evaluate predictivity of timepoint 2 *p* above-and-beyond timepoint 1 *p*, all timepoint two models (where timepoint 2 *p* factor was the outcome variable of interest) included timepoint 1 *p* factor as a covariate. Although timepoint 1 parental *p* significantly contributed to timepoint 2 child *p*, *g* and Grades failed to improve model fit.

| Cross-sectional deviance explained models | AIC |
| --- | --- |
| $p \sim s(g)$ | 73839.02 |
| $p \sim \text{Grades}$ | 73215.33 |
| $p \sim s(g) + s(\text{Parental } p)$ | 70125.89 |
| $p \sim \text{Grades} + s(\text{Parental } p)$ | 69531.53 |
| Timepoint two deviance explained models |  |
| $p_{t2} \sim s(p_{t1})$ | 33577.95 |
| $p_{t2} \sim s(p_{t1}) + s(g_{t1})$ | 33579.23 |
| $p_{t2} \sim s(p_{t1}) + \text{Grades}_{t1}$ | 33579.71 |
| $p_{t2} \sim s(p_{t1}) + s(g_{t1}) + s(\text{Parental } p_{t1})$ | 33521.32 |
| $p_{t2} \sim s(p_{t1}) + \text{Grades}_{t1} + s(\text{Parental } p_{t1})$ | 33520.14 |

## References

1. Friedrich, M. J. Depression Is the Leading Cause of Disability Around the World. JAMA 317, 1517 (2017).

2. Piao, J., Huang, Y., Han, C., Li, Y., Xu, Y., Liu, Y. & He, X. Alarming changes in the global burden of mental disorders in children and adolescents from 1990 to 2019: a systematic analysis for the Global Burden of Disease study. Eur Child Adolesc Psychiatry 31, 1827–1845 (2022).

3. Forbes, M. K., Neo, B., Nezami, O. M., Fried, E. I., Faure, K., Michelsen, B., Twose, M. & Dras, M. Elemental psychopathology: Distilling constituent symptoms and patterns of repetition in the diagnostic criteria of the DSM-5. Preprint at 10.31234/osf.io/u56p2 (2023).

4. Pan, Z., Park, C., Brietzke, E., Zuckerman, H., Rong, C., Mansur, R. B., Fus, D., Subramaniapillai, M., Lee, Y. & McIntyre, R. S. Cognitive impairment in major depressive disorder. CNS Spectrums 24, 22–29 (2019).

5. Lettau, J. The Impact of Children’s Academic Competencies and School Grades on their Life Satisfaction: What Really Matters? Child Ind Res 14, 2171–2195 (2021).

6. Wille, N., Bettge, S., Wittchen, H.-U., Ravens-Sieberer, U., & The BELLA study group. How impaired are children and adolescents by mental health problems? Results of the BELLA study. Eur Child Adolesc Psychiatry 17, 42–51 (2008).

7. Tozzi, L., Goldstein-Piekarski, A. N., Korgaonkar, M. S. & Williams, L. M. Connectivity of the Cognitive Control Network During Response Inhibition as a Predictive and Response Biomarker in Major Depression: Evidence From a Randomized Clinical Trial. Biol Psychiatry 87, 462–472 (2020).

8. Hack, L. M., Tozzi, L., Zenteno, S., Olmsted, A. M., Hilton, R., Jubeir, J., Korgaonkar, M. S., Schatzberg, A. F., Yesavage, J. A., O’Hara, R. & Williams, L. M. A Cognitive Biotype of Depression Linking Symptoms, Behavior Measures, Neural Circuits, and Differential Treatment Outcomes: A Prespecified Secondary Analysis of a Randomized Clinical Trial. JAMA Network Open 6, e2318411 (2023).

9. Morozova, A., Zorkina, Y., Abramova, O., Pavlova, O., Pavlov, K., Soloveva, K., Volkova, M., Alekseeva, P., Andryshchenko, A., Kostyuk, G., Gurina, O. & Chekhonin, V. Neurobiological Highlights of Cognitive Impairment in Psychiatric Disorders. International Journal of Molecular Sciences 23, 1217 (2022).

10. Williams, L. M. Precision psychiatry: a neural circuit taxonomy for depression and anxiety. Lancet Psychiatry 3, 472–480 (2016).

11. Mungas, D., Beckett, L., Harvey, D., Farias, S. T., Reed, B., Carmichael, O., Olichney, J., Miller, J. & DeCarli, C. Heterogeneity of Cognitive Trajectories in Diverse Older Person. Psychol Aging 25, 606–619 (2010).

12. Amalric, M. & Cantlon, J. F. Entropy, complexity, and maturity in children’s neural responses to naturalistic video lessons. Cortex 163, 14–25 (2023).

13. Camacho, M. C., Nielsen, A. N., Balser, D., Furtado, E., Steinberger, D. C., Fruchtman, L., Culver, J. P., Sylvester, C. M. & Barch, D. M. Large-scale encoding of emotion concepts becomes increasingly similar between individuals from childhood to adolescence. Nat Neurosci 26, 1256–1266 (2023).

14. Tansey, R., Graff, K., Rohr, C. S., Dimond, D., Ip, A., Yin, S., Dewey, D. & Bray, S. Functional MRI responses to naturalistic stimuli are increasingly typical across early childhood. Developmental Cognitive Neuroscience 62, 101268 (2023).

15. Cortina, M. A., Stein, A., Kahn, K., Hlungwani, T. M., Holmes, E. A. & Fazel, M. Cognitive styles and psychological functioning in rural South African school students: Understanding influences for risk and resilience in the face of chronic adversity. Journal of Adolescence 49, 38–46 (2016).

16. McLaughlin, K. A., Colich, N. L., Rodman, A. M. & Weissman, D. G. Mechanisms linking childhood trauma exposure and psychopathology: a transdiagnostic model of risk and resilience. BMC Medicine 18, 96 (2020).

17. Larsen, B. & Luna, B. Adolescence as a neurobiological critical period for the development of higher-order cognition. Neuroscience & Biobehavioral Reviews 94, 179–195 (2018).

18. Callaghan, B. L. & Tottenham, N. The Stress Acceleration Hypothesis: Effects of early-life adversity on emotion circuits and behavior. Curr Opin Behav Sci 7, 76–81 (2016).

19. Blakemore, S.-J. & Mills, K. L. Is Adolescence a Sensitive Period for Sociocultural Processing? Annual Review of Psychology 65, 187–207 (2014).

20. Blair, C. & Raver, C. C. Poverty, Stress, and Brain Development: New Directions for Prevention and Intervention. Academic Pediatrics 16, S30–S36 (2016).

21. Ridley, M., Rao, G., Schilbach, F. & Patel, V. Poverty, depression, and anxiety: Causal evidence and mechanisms. Science 370, eaay0214 (2020).

22. Fekadu, W., Mihiretu, A., Craig, T. K. J. & Fekadu, A. Multidimensional impact of severe mental illness on family members: systematic review. BMJ Open 9, e032391 (2019).

23. Stracke, M., Heinzl, M., Müller, A. D., Gilbert, K., Thorup, A. A. E., Paul, J. L. & Christiansen, H. Mental Health Is a Family Affair—Systematic Review and Meta-Analysis on the Associations between Mental Health Problems in Parents and Children during the COVID-19 Pandemic. Int J Environ Res Public Health 20, 4485 (2023).

24. Duarte, C. S., Monk, C., Weissman, M. M. & Posner, J. Intergenerational psychiatry: a new look at a powerful perspective. World Psychiatry 19, 175–176 (2020).

25. Yehuda, R. & Lehrner, A. Intergenerational transmission of trauma effects: putative role of epigenetic mechanisms. World Psychiatry 17, 243–257 (2018).

26. Belsky, J., Ruttle, P. L., Boyce, W. T., Armstrong, J. M. & Essex, M. J. Early adversity, elevated stress physiology, accelerated sexual maturation, and poor health in females. Developmental Psychology 51, 816–822 (2015).

27. Cowan, C. S. M. & Richardson, R. Early-life stress leads to sex-dependent changes in pubertal timing in rats that are reversed by a probiotic formulation. Dev Psychobiol 61, 679–687 (2019).

28. Casey, B., Jones, R. M. & Somerville, L. H. Braking and Accelerating of the Adolescent Brain. J Res Adolesc 21, 21–33 (2011).

29. McDermott, C. L., Hilton, K., Park, A. T., Tooley, U. A., Boroshok, A. L., Mupparapu, M., Scott, J. M., Bumann, E. E. & Mackey, A. P. Early life stress is associated with earlier emergence of permanent molars. Proceedings of the National Academy of Sciences 118, e2105304118 (2021).

30. Rakesh, D., Whittle, S., Sheridan, M. A. & McLaughlin, K. A. Childhood socioeconomic status and the pace of structural neurodevelopment: accelerated, delayed, or simply different? Trends in Cognitive Sciences 27, 833–851 (2023).

31. Rapee, R. M., Bőgels, S. M., van der Sluis, C. M., Craske, M. G. & Ollendick, T. Annual research review: conceptualising functional impairment in children and adolescents. J Child Psychol Psychiatry 53, 454–468 (2012).

32. Tordjman, S., Vaivre-Douret, L., Chokron, S. & Kermarrec, S. Les enfants à haut potentiel en difficulté : apports de la recherche clinique. L’Encéphale 44, 446–456 (2018).

33. González, C., Varela, J., Sánchez, P. A., Venegas, F. & De Tezanos-Pinto, P. Students’ Participation in School and its Relationship with Antisocial Behavior, Academic Performance and Adolescent Well-Being. Child Ind Res 14, 269–282 (2021).

34. Williams, C. M., Peyre, H., Labouret, G., Fassaya, J., García, A. G., Gauvrit, N. & Ramus, F. High intelligence is not associated with a greater propensity for mental health disorders. European Psychiatry 66, e3 (2023).

35. Snyder, H. R., Miyake, A. & Hankin, B. L. Advancing understanding of executive function impairments and psychopathology: bridging the gap between clinical and cognitive approaches. Frontiers in Psychology 6, (2015).

36. Snyder, H. R. Major depressive disorder is associated with broad impairments on neuropsychological measures of executive function: a meta-analysis and review. Psychol Bull 139, 81–132 (2013).

37. Abramovitch, A., Short, T. & Schweiger, A. The C Factor: Cognitive dysfunction as a transdiagnostic dimension in psychopathology. Clinical Psychology Review 86, 102007 (2021).

38. Mascio, A., Stewart, R., Botelle, R., Williams, M., Mirza, L., Patel, R., Pollak, T., Dobson, R. & Roberts, A. Cognitive Impairments in Schizophrenia: A Study in a Large Clinical Sample Using Natural Language Processing. Front Digit Health 3, 711941 (2021).

39. Kermarrec, S., Attinger, L., Guignard, J.-H. & Tordjman, S. Anxiety disorders in children with high intellectual potential. BJPsych Open 6, e70 (2020).

40. Karpinski, R. I., Kinase Kolb, A. M., Tetreault, N. A. & Borowski, T. B. High intelligence: A risk factor for psychological and physiological overexcitabilities. Intelligence 66, 8–23 (2018).

41. Smith, D. J., Anderson, J., Zammit, S., Meyer, T. D., Pell, J. P. & Mackay, D. Childhood IQ and risk of bipolar disorder in adulthood: prospective birth cohort study. BJPsych Open 1, 74–80 (2015).

42. Clarke, T.-K., Lupton, M. K., Fernandez-Pujals, A. M., Starr, J., Davies, G., Cox, S., Pattie, A., Liewald, D. C., Hall, L. S., MacIntyre, D. J., Smith, B. H., Hocking, L. J., Padmanabhan, S., Thomson, P. A., Hayward, C., Hansell, N. K., Montgomery, G. W., Medland, S. E., Martin, N. G., Wright, M. J., Porteous, D. J., Deary, I. J. & McIntosh, A. M. Common polygenic risk for autism spectrum disorder (ASD) is associated with cognitive ability in the general population. Mol Psychiatry 21, 419– 425 (2016).

43. Cullen, B., Nicholl, B. I., Mackay, D. F., Martin, D., Ul-Haq, Z., McIntosh, A., Gallacher, J., Deary, I. J., Pell, J. P., Evans, J. J. & Smith, D. J. Cognitive function and lifetime features of depression and bipolar disorder in a large population sample: Cross-sectional study of 143,828 UK Biobank participants. Eur Psychiatry 30, 950–958 (2015).

44. Shanmugan, S., Seidlitz, J., Cui, Z., Adebimpe, A., Bassett, D. S., Bertolero, M. A., Davatzikos, C., Fair, D. A., Gur, R. E., Gur, R. C., Larsen, B., Li, H., Pines, A., Raznahan, A., Roalf, D. R., Shinohara, R. T., Vogel, J., Wolf, D. H., Fan, Y., Alexander-Bloch, A. & Satterthwaite, T. D. Sex differences in the functional topography of association networks in youth. Proceedings of the National Academy of Sciences 119, e2110416119 (2022).

45. Bethlehem, R. a. I., Seidlitz, J., White, S. R., Vogel, J. W., Anderson, K. M., Adamson, C., Adler, S., Alexander-Bloch, A. F., et al. Brain charts for the human lifespan. Nature 604, 525–533 (2022).

46. Pines, A. R., Larsen, B., Cui, Z., Sydnor, V. J., Bertolero, M. A., Adebimpe, A., Alexander-Bloch, A. F., Davatzikos, C., Fair, D. A., Gur, R. C., Gur, R. E., Li, H., Milham, M. P., Moore, T. M., Murtha, K., Parkes, L., Thompson-Schill, S. L., Shanmugan, S., Shinohara, R. T., Weinstein, S. M., Bassett, D. S., Fan, Y. & Satterthwaite, T. D. Dissociable multi-scale patterns of development in personalized brain networks. Nat Commun 13, 2647 (2022).

47. Larsen, B., Bourque, J., Moore, T. M., Adebimpe, A., Calkins, M. E., Elliott, M. A., Gur, R. C., Gur, R. E., Moberg, P. J., Roalf, D. R., Ruparel, K., Turetsky, B. I., Vandekar, S. N., Wolf, D. H., Shinohara, R. T. & Satterthwaite, T. D. Longitudinal Development of Brain Iron Is Linked to Cognition in Youth. J Neurosci 40, 1810– 1818 (2020).

48. Larsen, B., Baller, E. B., Boucher, A. A., Calkins, M. E., Laney, N., Moore, T. M., Roalf, D. R., Ruparel, K., Gur, R. C., Gur, R. E., Georgieff, M. K. & Satterthwaite, T. D. Development of Iron Status Measures during Youth: Associations with Sex, Neighborhood Socioeconomic Status, Cognitive Performance, and Brain Structure. Am J Clin Nutr 118, 121–131 (2023).

49. Hastie, T. & Tibshirani, R. Generalized additive models for medical research. Stat Methods Med Res 4, 187–196 (1995).

50. Wood, S. N. Fast stable restricted maximum likelihood and marginal likelihood estimation of semiparametric generalized linear models. Journal of the Royal Statistical Society. Series B (Statistical Methodology) 73, 3–36 (2011).

51. Garavan, H., Bartsch, H., Conway, K., Decastro, A., Goldstein, R. Z., Heeringa, S., Jernigan, T., Potter, A., Thompson, W. & Zahs, D. Recruiting the ABCD sample: Design considerations and procedures. Dev Cogn Neurosci 32, 16–22 (2018).

52. Jernigan, T. L., Brown, S. A. & Dowling, G. J. The Adolescent Brain Cognitive Development Study. J Res Adolesc 28, 154–156 (2018).

53. U.S. Census Bureau QuickFacts: United States. https://www.census.gov/quickfacts/fact/table/US/PST045222.

54. United Nations Guide for Minorities. OHCHR https://www.ohchr.org/en/minorities/united-nations-guide-minorities.

55. Thompson, W. K., Barch, D. M., Bjork, J. M., Gonzalez, R., Nagel, B. J., Nixon, S. J. & Luciana, M. The structure of cognition in 9 and 10 year-old children and associations with problem behaviors: Findings from the ABCD study’s baseline neurocognitive battery. Dev Cogn Neurosci 36, 100606 (2019).

56. Warne, R. T. & Burningham, C. Spearman’s g found in 31 non-Western nations: Strong evidence that g is a universal phenomenon. Psychol Bull 145, 237–272 (2019).

57. Johnson, W., Bouchard, T. J., Krueger, R. F., McGue, M. & Gottesman, I. I. Just one g: consistent results from three test batteries. Intelligence 32, 95–107 (2004).

58. Achenbach, T. M. & Rescorla, L. Manual for the ASEBA school-age forms & profiles: an integrated system of multi-informant assessment. (ASEBA, 2001).

59. Zelenina, M., Pine, D., Stringaris, A. & Nielson, D. Validation of CBCL depression scores of adolescents in three independent datasets. Preprint at 10.31234/osf.io/z956k (2023).

60. Michelini, G., Barch, D. M., Tian, Y., Watson, D., Klein, D. N. & Kotov, R. Delineating and validating higher-order dimensions of psychopathology in the Adolescent Brain Cognitive Development (ABCD) study. Translational Psychiatry 9, 1–15 (2019).

61. Feczko, E., Conan, G., Marek, S., Tervo-Clemmens, B., Cordova, M., Doyle, O., Earl, E., Perrone, A., Sturgeon, D., Klein, R., Harman, G., Kilamovich, D., Hermosillo, R., Miranda-Dominguez, O., Adebimpe, A., Bertolero, M., Cieslak, M., Covitz, S., Hendrickson, T., Juliano, A. C., Snider, K., Moore, L. A., Uriartel, J., Graham, A. M., Calabro, F., Rosenberg, M. D., Rapuano, K. M., Casey, B. J., Watts, R., Hagler, D., Thompson, W. K., Nichols, T. E., Hoffman, E., Luna, B., Garavan, H., Satterthwaite, T. D., Ewing, S. F., Nagel, B., Dosenbach, N. U. F. & Fair, D. A. Adolescent Brain Cognitive Development (ABCD) Community MRI Collection and Utilities. 2021.07.09.451638 Preprint at 10.1101/2021.07.09.451638 (2021).

62. Stumm, S. von, Malanchini, M. & Fisher, H. L. The developmental interplay between the p-factor of psychopathology and the g-factor of intelligence from age 7 through 16 years. Development and Psychopathology 1–10 (2023) doi:10.1017/S095457942300069X.

63. Romer, A. L. & Pizzagalli, D. A. Is executive dysfunction a risk marker or consequence of psychopathology? A test of executive function as a prospective predictor and outcome of general psychopathology in the adolescent brain cognitive development study®. Developmental Cognitive Neuroscience 51, 100994 (2021).

64. Kenny, D. A. Cross-lagged panel correlation: A test for spuriousness. Psychological Bulletin 82, 887–903 (1975).

65. Pines, A. R., Cieslak, M., Larsen, B., Baum, G. L., Cook, P. A., Adebimpe, A., Dávila, D. G., Elliott, M. A., Jirsaraie, R., Murtha, K., Oathes, D. J., Piiwaa, K., Rosen, A. F. G., Rush, S., Shinohara, R. T., Bassett, D. S., Roalf, D. R. & Satterthwaite, T. D. Leveraging multi-shell diffusion for studies of brain development in youth and young adulthood. Developmental Cognitive Neuroscience 43, 100788 (2020).

66. Klapwijk, E. T., van den Bos, W., Tamnes, C. K., Raschle, N. M. & Mills, K. L. Opportunities for increased reproducibility and replicability of developmental neuroimaging. Developmental Cognitive Neuroscience 47, 100902 (2021).

67. Cardenas-Iniguez, C., Moore, T. M., Kaczkurkin, A. N., Meyer, F. A. C., Satterthwaite, T. D., Fair, D. A., White, T., Blok, E., Applegate, B., Thompson, L. M., Rosenberg, M. D., Hedeker, D., Berman, M. G. & Lahey, B. B. Direct and Indirect Associations of Widespread Individual Differences in Brain White Matter Microstructure With Executive Functioning and General and Specific Dimensions of Psychopathology in Children. Biological Psychiatry: Cognitive Neuroscience and Neuroimaging 7, 362–375 (2022).

68. Wiegand-Grefe, S., Sell, M., Filter, B. & Plass-Christl, A. Family Functioning and Psychological Health of Children with Mentally Ill Parents. International Journal of Environmental Research and Public Health 16, (2019).

69. Mensah, F. K. & Kiernan, K. E. Parents’ mental health and children’s cognitive and social development. Soc Psychiat Epidemiol 45, 1023–1035 (2010).

70. Tooley, U. A., Bassett, D. S. & Mackey, A. P. Environmental influences on the pace of brain development. Nature Reviews Neuroscience 1–13 (2021) doi:10.1038/s41583-021-00457-5.

71. Rechlin, R. K., Splinter, T. F. L., Hodges, T. E., Albert, A. Y. & Galea, L. A. M. An analysis of neuroscience and psychiatry papers published from 2009 and 2019 outlines opportunities for increasing discovery of sex differences. Nat Commun 13, 2137 (2022).

72. Lai, A. G. & Chang, W. H. There is no health without mental health: Challenges ignored and lessons learned. Clin Transl Med 12, e897 (2022).

73. De Los Reyes, A., Augenstein, T. M., Wang, M., Thomas, S. A., Drabick, D. A. G., Burgers, D. E. & Rabinowitz, J. The validity of the multi-informant approach to assessing child and adolescent mental health. Psychol Bull 141, 858–900 (2015).

74. Hughes, E. K. & Gullone, E. Discrepancies between adolescent, mother, and father reports of adolescent internalizing symptom levels and their association with parent symptoms. Journal of Clinical Psychology 66, 978–995 (2010).

75. Reedtz, C., van Doesum, K., Signorini, G., Lauritzen, C., van Amelsvoort, T., van Santvoort, F., Young, A. H., Conus, P., Musil, R., Schulze, T., Berk, M., Stringaris, A., Piché, G. & de Girolamo, G. Promotion of Wellbeing for Children of Parents With Mental Illness: A Model Protocol for Research and Intervention. Front Psychiatry 10, 606 (2019).

76. Morawska, A. & Sanders, M. An evaluation of a behavioural parenting intervention for parents of gifted children. Behaviour Research and Therapy 47, 463–470 (2009).

77. Lundahl, B., Risser, H. J. & Lovejoy, M. C. A meta-analysis of parent training: moderators and follow-up effects. Clin Psychol Rev 26, 86–104 (2006).

78. Weissman, M. M. Children of Depressed Parents—A Public Health Opportunity. JAMA Psychiatry 73, 197–198 (2016).

79. Crosby, D., Bossutt, P., Brockehurst, P., Chamberlain, C., Dive, C., Holmes, C., Isaacs, J., Kennedy, R., Matthews, F., Parmer, M., Pearce, J., Westhead, D., Whittaker, J. & Holgate, S. The MRC Framework for the Development, Design and Analysis of Stratified Medicine Research: Enabling Stratified, Precision and Personalised Medicine. https://eprints.ncl.ac.uk (2017).

80. Falconnier, L. Socioeconomic Status in the Treatment of Depression. American Journal of Orthopsychiatry 79, 148–158 (2009).

81. Alegria, M., Falgas-Bague, I. & Fong, H. Engagement of ethnic minorities in mental health care. World Psychiatry 19, 35–36 (2020).

82. Zeira, A. Mental Health Challenges Related to Neoliberal Capitalism in the United States. Community Ment Health J 58, 205–212 (2022).

83. Maj, M. Beyond diagnosis in psychiatric practice. Annals of General Psychiatry 19, 27 (2020).

84. All of Us Research Program | National Institutes of Health (NIH). All of Us Research Program | NIH https://allofus.nih.gov/future-health-begins-all-us (2020).

85. Ge, J., Yang, G., Han, M., Zhou, S., Men, W., Qin, L., Lyu, B., Li, H., Wang, H., Rao, H., Cui, Z., Liu, H., Zuo, X.-N. & Gao, J.-H. Increasing diversity in connectomics with the Chinese Human Connectome Project. Nat Neurosci 26, 163–172 (2023).

86. Sharma, E., Vaidya, N., Iyengar, U., Zhang, Y., Holla, B., Purushottam, M., Chakrabarti, A., Fernandes, G. S., Heron, J., Hickman, M., Desrivieres, S., Kartik, K., Jacob, P., Rangaswamy, M., Bharath, R. D., Barker, G., Orfanos, D. P., Ahuja, C., Murthy, P., Jain, S., Varghese, M., Jayarajan, D., Kumar, K., Thennarasu, K., Basu, D., Subodh, B. N., Kuriyan, R., Kurpad, S. S., Kalyanram, K., Krishnaveni, G., Krishna, M., Singh, R. L., Singh, L. R., Kalyanram, K., Toledano, M., Schumann, G., Benegal, V., & The cVEDA Consortium. Consortium on Vulnerability to Externalizing Disorders and Addictions (cVEDA): A developmental cohort study protocol. BMC Psychiatry 20, 2 (2020).

87. Fan, X.-R., Wang, Y.-S., Chang, D., Yang, N., Rong, M.-J., Zhang, Z., He, Y., Hou, X., Zhou, Q., Gong, Z.-Q., Cao, L.-Z., Dong, H.-M., Nie, J.-J., Chen, L.-Z., Zhang, Q., Zhang, J.-X., Zhang, L., Li, H.-J., Bao, M., Chen, A., Chen, J., Chen, X., Ding, J., Dong, X., Du, Y., Feng, C., Feng, T., Fu, X., Ge, L.-K., Hong, B., Hu, X., Huang, W., Jiang, C., Li, L., Li, Q., Li, S., Liu, X., Mo, F., Qiu, J., Su, X.-Q., Wei, G.-X., Wu, Y., Xia, H., Yan, C.-G., Yan, Z.-X., Yang, X., Zhang, W., Zhao, K., Zhu, L. & Zuo, X.-N. A longitudinal resource for population neuroscience of school-age children and adolescents in China. Sci Data 10, 545 (2023).

88. Elyounssi, S., Kunitoki, K., Clauss, J. A., Laurent, E., Kane, K., Hughes, D. E., Hopkinson, C. E., Bazer, O., Sussman, R. F., Doyle, A. E., Lee, H., Tervo-Clemmens, B., Eryilmaz, H., Gollub, R. L., Barch, D. M., Satterthwaite, T. D., Dowling, K. F. & Roffman, J. L. Uncovering and mitigating bias in large, automated MRI analyses of brain development. 2023.02.28.530498 Preprint at 10.1101/2023.02.28.530498 (2023).

89. Ricard, J. A., Parker, T. C., Dhamala, E., Kwasa, J., Allsop, A. & Holmes, A. J. Confronting racially exclusionary practices in the acquisition and analyses of neuroimaging data. Nat Neurosci 26, 4–11 (2023).

90. Berger, C., Lisboa, C., Cuadros, O. & de Tezanos-Pinto, P. Adolescent Peer Relations and Socioemotional Development in Latin America: Translating International Theory into Local Research. New Directions for Child and Adolescent Development 2016, 45–58 (2016).

91. Whitmore, L. B. & Mills, K. L. Co-creating developmental science. Infant and Child Development 31, e2273 (2022).

